# Modeling geographical accessibility and inequalities to childbirth services in the *Grand Nokoué* metropolitan area, Benin

**DOI:** 10.1101/2025.05.15.25327676

**Authors:** Yiséché SC Hounménou, Elias Martinien Avahoundjè, Aline Semaan, Christian Mahugnon Agossou, Christelle Boyi Hounsou, Mena Komi Agbodjavou, Giulia Scarpa, Ange D Dossou, Thierry Lawale, Romuald Bothon, Justin Lewis Denakpo, Lenka Beňová, Jean-Paul Dossou, Peter M Macharia

**Affiliations:** Centre de recherche en reproduction humaine et en démographie (Cerrhud), Cotonou, Bénin; Department of Public Health, Institute of Tropical Medicine, Antwerp, Belgium; Ministry of Health, Cotonou, Bénin; Centre National Hospitalier Universitaire Hubert K. Maga de Cotonou; Faculty of Epidemiology and Population Health, London School of Hygiene & Tropical Medicine, London, UK; Population & Health Impact Surveillance Group, Kenya Medical Research Institute-Wellcome Trust Research Programme, Nairobi, Kenya

**Keywords:** Spatial accessibility, travel time, maternal health, urban areas, relative wealth, inequities, Benin

## Abstract

Timely access to comprehensive, high-quality emergency obstetric and neonatal care can prevent maternal and neonatal mortality, but remains challenging in Benin. We examine geographic accessibility to childbirth care in *Grand Nokoué*, the largest conurbation with five urban areas in Benin.

We gathered data on boundaries, health facilities, road network, elevation, land cover, relative wealth, urbanicity and geo-traced travel speeds in *Grand Nokoué*. We modelled travel times to health facility offering childbirth care (stratified by level and sector) using the least-cost path algorithm, based on slowest, average and fastest travel speeds. We estimated the percentage of women of childbearing age (WoCBA) within 30, 60, and 120 minutes of the nearest facility by subnational area used for decision making. We explored inequalities in travel time by wealth quintiles and urbanicity gradient.

Travel time was 8-minutes (31-minutes for hospital) at average speed and 24 minutes (106-minutes for hospital) at slowest speed to nearest facility. All and 71.6% of WoCBA live within 30 minutes of a health facility and hospital, respectively at average speeds. This decreases to 84.9% and 22.9%, respectively at slowest speed. There were substantial variations in travel time and geographic coverage at subnational units. The poorest travelled 5 times longer than the richest quintile, while travel time was shorter in the core urban relative to peri-urban.

Travel time to childbirth care in *Grand Nokoué*, varies by wealth and residence area type and is longer at the slowest speeds. Targeting peri-urban areas and poorest WoCBA with longer travel time will reduce inequities.

## Introduction

The majority (70%) of maternal deaths globally occur in sub-Saharan Africa (SSA), where the maternal mortality ratio (MMR) was 545 per 100,000 livebirths in 2020 [1]. It is unlikely that the Sustainable Development Goal target 3.1, to reduce MMR to less than 70 per 100,000 livebirths, by 2030 will be achieved in SSA [1]. Furthermore, 44% of the estimated 2 million stillbirths worldwide occurred in SSA in 2019 [2].

Half of maternal deaths and three-quarters of intrapartum stillbirths are preventable if timely access to high-quality comprehensive emergency obstetric and newborn care (CEmONC) provided by skilled personnel is available [3]. However, timely geographical access to CEmONC, mostly provided in hospitals, remains a problem in most SSA countries [4,5]. In 2018, 28% of women of childbearing age (WoCBA-15-49 years) in SSA resided more than 2-hours away from the nearest hospital [4]. The World Health Organization (WHO) recommends that CEmONC be available within 2–3 hours travel time [6]. However, this window might be too long because delays of even 10-30 minutes are associated with stillbirths and maternal mortality [7].

Urban areas are unique when it comes to maternal and newborn health [8]. Historically, urban areas in SSA offered several advantages including better infrastructure, educational and healthcare facilities, and better sanitation compared to rural areas [9,10]. Urban residents have had relatively better health outcomes than rural residents, the so-called “urban advantage” [9–11]. However, this advantage is diminishing or becoming less apparent in some urban areas in both neonatal and adult mortality [12–14].

SSA is also experiencing the fastest rates of urbanisation globally [15]. Approximately 90% of the additional 2.5 billion urban residents will reside in Africa and Asia by 2050 [15], and the world’s biggest cities will be concentrated in Africa by 2100 [16]. Further, maternal healthcare service provision in urban areas of SSA has not matched the current and projected degree of urbanization [8,9,11]. This situation exacerbates challenges related to maternal and newborn health due to several factors, including long travel times to healthcare despite short distances partly due to traffic, haphazardly built environments, informal settlements’ growth, urban poverty, air pollution and environmental degradation [8,13,17].

Benin, a country in West Africa, has a high MMR of 523 per 100,000 livebirths [1]. This is despite various national governmental policies, strategies and interventions over the past decades to promote maternal health [18]. A closer and disaggregated evaluation of accessibility to and quality of care is therefore necessary. Six in ten WoCBA face problems accessing healthcare, related to getting permission, going alone, getting money for medical prescriptions or distance to health facility [19]. Among these, spatial access and place of residence are significant barriers to timely access [19–21]; long travel time negatively affects use of antenatal care, facility-based childbirth care, and skilled birth attendance in Benin [20].

Several studies have assessed geographical accessibility to care either at the national level or included Benin as part of regional (SSA) analysis [4,5,20,22–25]. Most of these studies used a database assembled between 2012 and 2018 of public-sector facilities only [26], thus not capturing the current landscape of healthcare service provision. Private sector providers (non-governmental organizations-NGO, faith-based Organization-FBO and for-profit facilities) were excluded, although they provide two-fifths of reproductive and maternal healthcare in SSA; this is likely to be higher in urban settings. Importantly, the approaches used to derive travel time in some of these analyses did not consider transport barriers like topography and traffic congestion, specifically challenging in urban contexts. Recent studies have highlighted the value of context-specific approaches to estimating geographic accessibility in urban areas [27,28].

This paper aims to estimate population-level average travel time to facility-based childbirth care in *Grand Nokoué* metropolitan area, Benin’s most urbanized and economically dynamic region[29]. We assess wealth and urbanicity related inequalities in geographic accessibility and estimate the percentage of women who live within 30, 60 and 120 minutes of the nearest facility providing childbirth care.

## Methods

### Setting: Benin and *Grand Nokoué* metropolitan area

Benin is divided into 12 main administrative subdivisions (*départements*) and further into 77 communes. Benin’s population was 14.1 million in 2023 and is projected at 24.4 million by 2050 [30]. The share of the population living in urban areas increased from 9.3% in 1960 to 50.1% in 2023 and projected at 65.5% by 2050 [31]. Majority of the urban population (80%) lives in the country’s southern coastal area – the *Grand Nokoué*, government designated metropolitan area (**Fig. 1**). The *Grand Nokoué* consists of five administrative communes: Cotonou (economic capital), Porto-Novo (administrative capital), Ouidah (tourist center), Abomey-Calavi (industrial hub), and *Sèmè-kpodji* (business and industrial center). Each commune is subdivided into several *arrondissements* (total of 43 in the *Grand Nokoué)*. In terms of service delivery the *Grand Nokoué* consists of seven health zones (**Supplementary material Fig. S1**), which are part of three administrative regions (*Département* of Atlantique, Littoral, and *Ouémé*), **Supplementary material Fig. S2**. The *Grand Nokoué* is densely populated, with2.38 million inhabitants in 2020 [29] (about 20% of Benin’s population) in an area of 1,444 Km^2^. This population is projected to reach over 3 million by 2030 [29]. The context of health service delivery is presented in **Supplementary material section 1.**

**Fig. 1.**
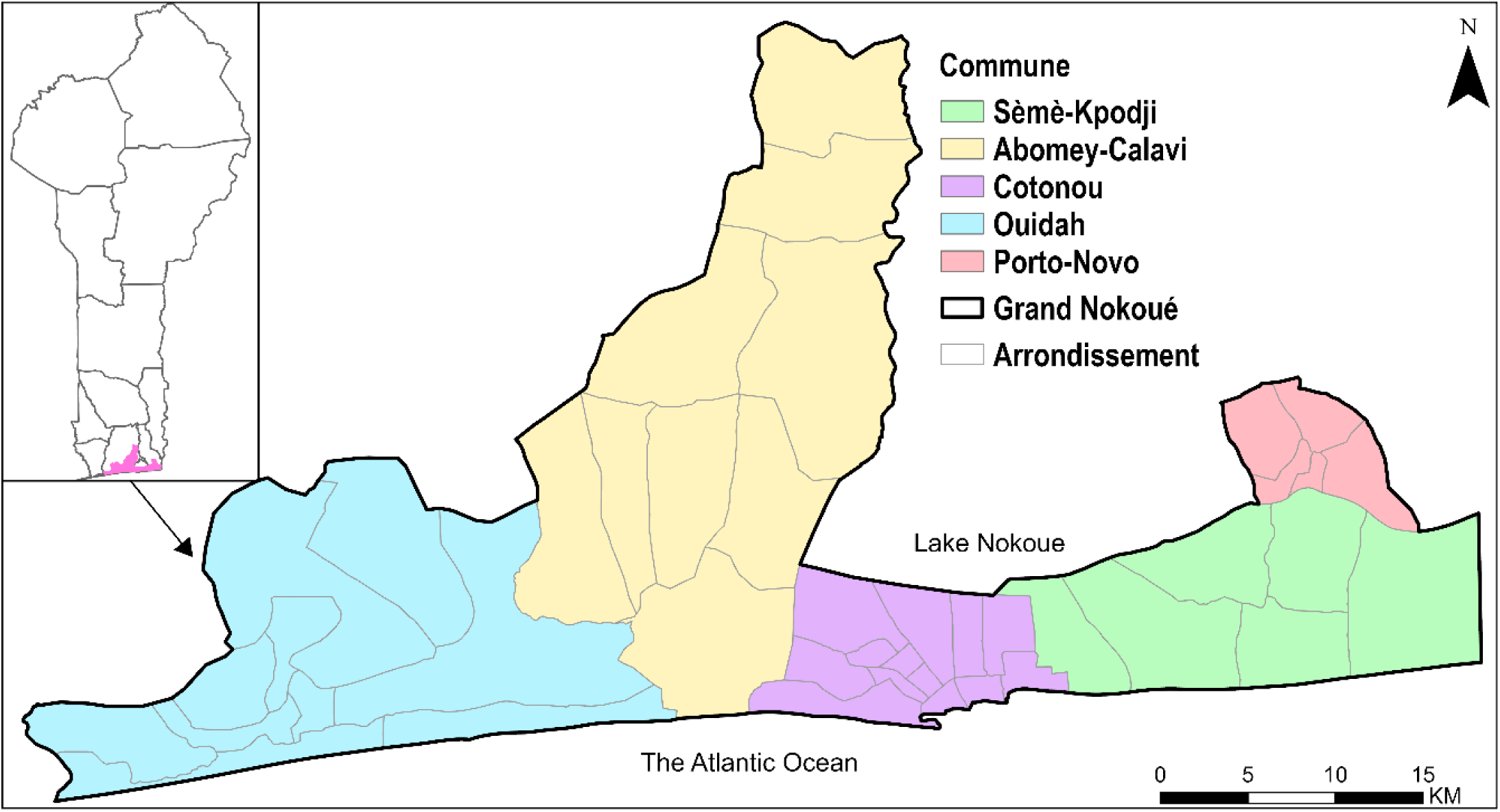
The *Grand Nokoué* metropolitan area, Benin comprising of five communes, and 43 *arrondissements*. Maps of three *Départements* and seven health zones are shown in **supplementary Figures S1 and S2**.

### Overview of methods

We followed four steps. First, we assembled databases of health facilities, factors affecting travel (road network, land use, and digital elevation model-DEM), gridded surfaces showing population distribution of WoCBA, relative wealth index (RWI), and urbanicity gradient. Second, we geo-traced travel speeds across different road types in the *Grand Nokoué* to estimate realistic travel speeds. Third, we used a geospatial framework anchored on a least cost path algorithm to compute travel time to the nearest heath facility (by level and sector) for different travel speed scenarios. Finally, we spatially overlaid the travel estimates with RWI, urbanicity gradient and population distribution of WoCBA to assess inequalities.

### Data

Detailed data assembly process is described in the **Supplementary Material**. Briefly, we assembled a geocoded database of health facilities offering childbirth care in 2023 from Benin’s health information system augmented by a list of health facilities from the Ministry of Health. The road network (**Supplementary material Fig. S3**) was downloaded from OpenStreetMap (OSM). Land use/cover **(Supplementary material Fig. S3)** from Sentinel-2 imagery was used to represent areas between roads. The DEM from Shuttle Radar Topographic Mission (**Supplementary material Fig. S5**) was used to adjust walking speeds [32]. To assess equity, we used gridded surfaces of relative wealth index (RWI) (**Supplementary material Fig. S6)**, a proxy for relative standard of living and urbanicity gradient (**Supplementary material Fig S7**) showing settlements in the continuum of a hamlet to a mega-city. High-resolution gridded datasets of WoCBA **(Supplementary material Fig. S6)** were available from Worldpop. Finally, we geo-traced individual motorized journeys (trajectories) within in *Grand Nokoué* using tablets or smartphones to estimate motorized travel speeds for different road types in the *Grand Nokoué* based on a previous approach [27], **Supplementary material section 3**.

### Modelling travel time

We modelled travel time to the nearest facility offering childbirth care based on the least cost path algorithm. The least cost path was defined as the path that would take the least amount of time to reach a facility while accounting for the mode of transport, speed, road network, topography, and travel barriers. Travel time was estimated from 30m square grids to all facilities and separately, to all hospitals and a subset of facilities by level in the public sector. For each of four categories of health facilities, three models were estimated using different travel speeds; the slowest and fastest speed to provide a continuum of the worst and the best travel scenarios, as well as the average speed. The least-cost path algorithm was implemented in AccessMod software version 5.8.0, a WHO tool used to model spatial accessibility and coverage [33]. Further details are presented in **Supplementary material section 3**.

We used the zonal statistics function of ArcGIS Pro v 3.3.1 (ESRI, Redlands, CA, USA) to summarize the average travel time across the *Grand Nokoué* Metropolitan areas and sub-nationally for each health zone, commune and *arrondissement*.

### Travel time and spatial inequalities

The resulting gridded surface of travel time constrained within populated areas was spatially overlaid with geospatial layers of population distribution of WoCBA, urbanicity gradient, and RWI to assess inequalities. First, we estimated the proportion of WoCBA within specific travel time thresholds (30 minutes, 60 minutes, and 120 minutes) to reflect the geographic coverage and aggregated sub-nationally. These thresholds are widely used as benchmarks for policy and research on spatial access to services in SSA [4,5].

Second, we matched each travel time grid with the nearest estimate of RWI. Equiplots were then produced to summarize the variation of travel time across the RWI quintiles for the study area and each of the three *départements* using *ggplot2* package in R software (version 4.4.0). The disaggregation was done at the departmental level given the small sample of RWI data points in the entire study area. Finally, we extracted the average travel time within each urbanicity band ranging from city (large settlement) to very low density, almost uninhabited areas.

## Results

### Health facilities

The final geocoded database of health facilities had 292 facilities offering childbirth care within the metropolitan area (**Fig 2**). The majority of the facilities were in the private-for-profit sector (71%) and the rest were in the public sector – which are managed by either the government (24%) or FBOs (5%). There were 13 hospitals (10 in the public sector); the rest were either health centres (75; 69 in public sector), medical centres (48; 3 in public sector), or clinics (156; 2 in public sector).

**Fig. 2.**
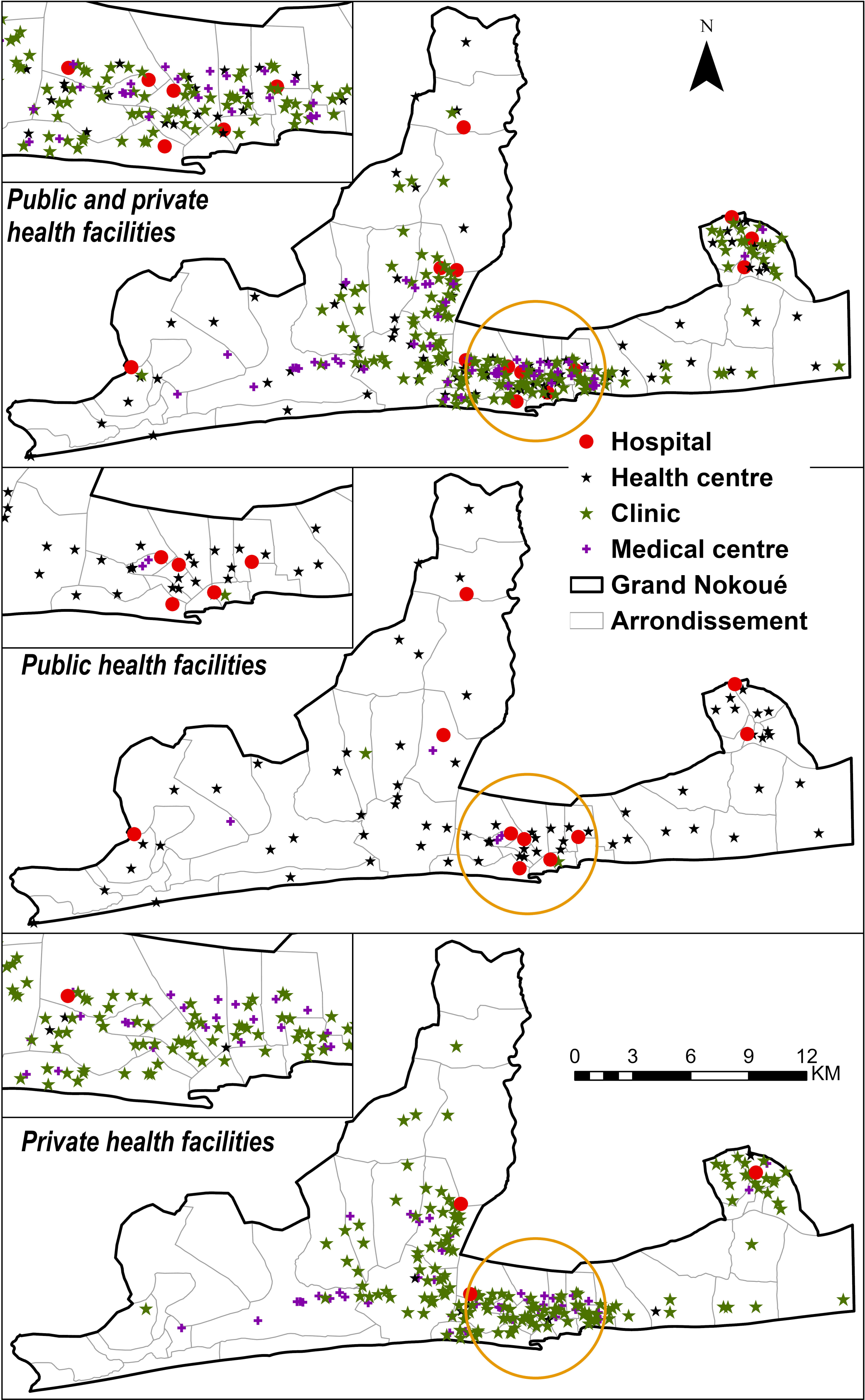
Locations of all 292 health facilities (29% in the public sector (24% managed by the government and 5% by confessional) and 71% in the private sector) offering childbirth care in the *Grand Nokoué* metropolitan area in Benin in 2023, by level

### Travel speed

The average travel speeds varied by road type (**Table 1**). Major roads, such as trunk roads (20.4km/hr) and primary roads (18.0km/hr), were characterized by relatively faster average speeds, whereas residential roads (12.6km/hr) and service roads (9.6 km/hr) were slower. The fastest travel speeds ranged from 25.5 km/hr to 38.0 km/hr and the slowest speeds ranged from 0.8km/hr to 6.2 km/hr (**Table 1**).

**Table 1.** Speeds by road type and land use/cover classes used to compute travel time to childbirth care facilities in *Grand Nokoué* metropolitan area, Benin.

| Road type (count of segments) | Description | Speed (km/hr) |  |  |  |
| --- | --- | --- | --- | --- | --- |
|  | Road Class | Mean | Min | Max | sd |
| Trunk (49) | Most important roads in a country | 20.4 | 6.2 | 38.0 | 8.9 |
| Primary (123) | Links larger towns | 18.0 | 3.1 | 38.0 | 9.0 |
| Secondary (57) | Links towns. | 15.1 | 4.4 | 30.0 | 5.8 |
| Tertiary (100) | Links smaller towns and villages | 15.3 | 4.1 | 33.3 | 8.5 |
| Residential (241) | Serve as an access to housing | 12.6 | 0.84 | 38.0 | 7.0 |
| Service (52) | Access to, or within business park, car park, etc. | 9.6 | 4.2 | 25.5 | 4.1 |
| Unclassified (35) | Links villages and hamlets (least important). Includes all tracks | 17.1 | 6.2 | 32.0 | 7.8 |
| Others (0) | Included foot way, cycleway, steps, living street.- walking trails- similar to bare ground and build up area | 5.0 | 4.0 | 6.0 | - |
| **Land use/cover** |  |  |  |  |  |
| Water | Water predominantly present throughout the year | 0 | 0 | 0 | - |
| Trees | Significant clustering of tall dense vegetation | 2.5 | 2.0 | 3.0 | - |
| Flooded vegetation | Vegetated areas with intermixing of water throughout a majority of the year | 0.1 | 0.08 | 0.12 | - |
| Crops | Human planted cereals, grasses, and crops not at tree height | 4.0 | 3.6 | 4.8 | - |
| Built Area | Human made structures e.g. roads and houses | 5.0 | 4.0 | 6.0 | - |
| Rangeland | Open areas covered with little to no taller vegetation | 4.5 | 4.05 | 5.4 | - |
| Bare ground | Areas with rock, soil, desert sand, with very sparse to no vegetation for the entire year | 5.0 | 4.0 | 6.0 | - |

Min- minimum speed, max-maximum speed, sd- standard deviation.

### Estimates of travel time

The average travel time to the nearest facility offering childbirth care in *Grand Nokoué* was 8 minutes in the average speed scenario. However, this increased to 24 minutes during the slowest speed scenario and reduced to 4 minutes in the fastest speed scenario. Average travel time to the nearest *public* facility offering childbirth care was 10 minutes, ranging from 5 to 32 minutes by scenario. The high-resolution maps are shown in **supplementary materials Figures S8 and S9.**

There were large differences in travel time between arrondissements (**Fig 3)**. The average travel time to nearest facility varied from 2 minutes (range 1 to 7 minutes) in *7ème arrondissement* in Cotonou to 38 minutes (27–65) in *Djègbadji arrondissement* (Ouidah). The range was similar when considering travel to public facilities only, from 3 minutes (2–11) in *10^ème^ arrondissement* in Cotonou to 38 (27–66) in *Djègbadji* (Ouidah) (**Fig 3**). Overall, *arrondissements* within Cotonou and Porto-Novo had shorter travel times compared to those in Ouidah (**Fig 3**). For example, the mean travel time to public facilities was 5 (3-19) and 6 (3-21) minutes in the communes of Cotonou and Porto-Novo, respectively, while Ouidah and Abomey-Calavi recorded 13 (7-35) and 11 (5-36) minutes, respectively (**Fig 3**).

**Fig. 3.**
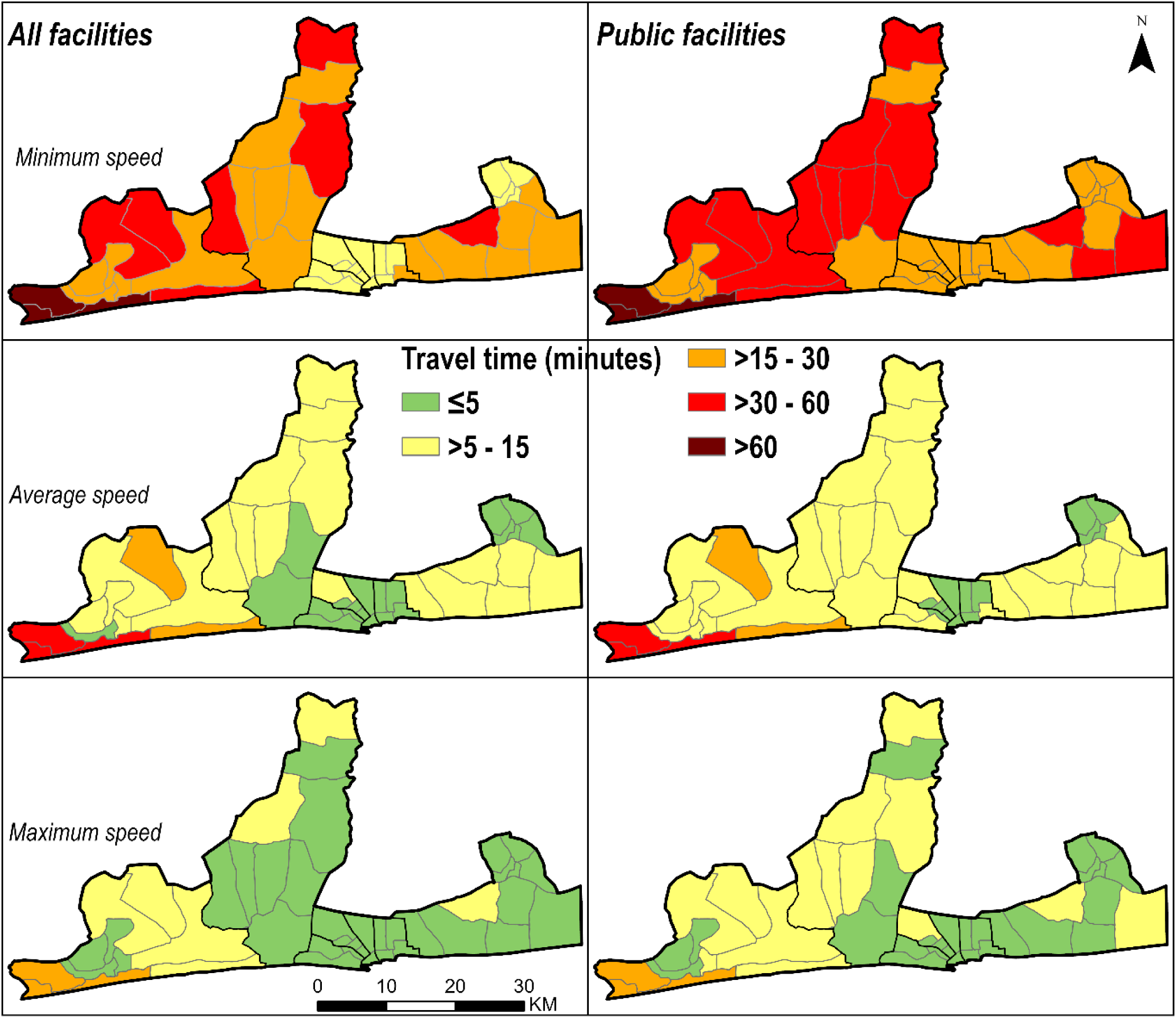
A*r*rondissement-level travel time to facilities offering childbirth care, by travel speed scenario (minimum, average and maximum) in *Grand Nokoué* metropolitan area, Benin. The dark boundary lines represent the seven health zones. The high resolution maps (raster cells at 30m) are shown in **Supplementary Figures S8 and S9**).

When we restricted accessibility to *hospitals* only, the average travel time was 31 minutes, ranging from 14 to 106 minutes depending on speed scenario. Models of access to *public hospitals* resulted in similar estimates (33 minutes, range 15-106). Women from communes of Cotonou and Porto-Novo had shorter travel times to hospitals, and from Abomey-Calavi and Ouidah longer. Across *arrondissements*, travel time to hospitals ranged from 4 (2-20) minutes in *8^ème^ arrondissement* of Cotonou to 68 (40-157) minutes in Djègbadji *arrondissement* (Ouidah) (**Fig 4**). In the slowest speed scenario, women from 10 out of the 43 *arrondissements* had average travel times to hospitals above 120-minutes (**Fig 4**).

**Fig. 4.**
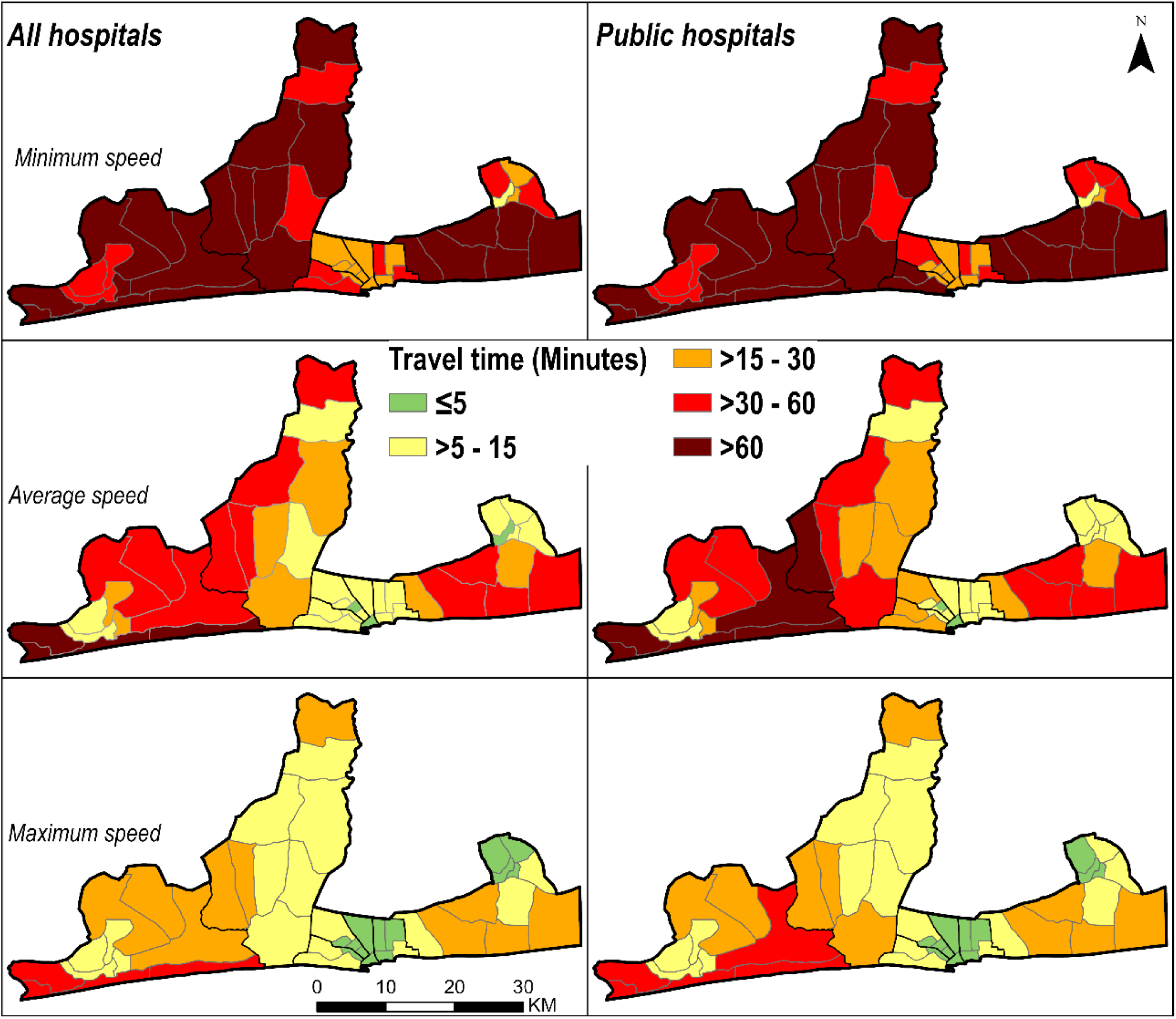
A*r*rondissement-level travel time to childbirth care in hospitals by travel speed scenario (minimum, average and maximum) in *Grand Nokoué* metropolitan area, Benin ( reposition legend to show it’s for all)

### Geographic coverage

In the metropolitan area, almost all WoCBA (99.5%) lived within 30 minutes of any facility offering childbirth care, considering average speed (99.5% for public facilities). However, in the slowest speed scenario, this reduced to 84.7% for all facilities and 64.5% for public facilities (**Table 2**). At the health zone level, the coverage was more heterogenous, especially at minimum speeds (**Table 2**). More variation was observed across the *arrondissement* level, ranging from 11.6% to 100% at the 30-minute threshold considering slowest travel speeds to public facilities. The percentage of WoCBA within 30-, 60- and 120-minute travel time thresholds are shown by commune and *arrondissement* in **Supplementary material Tables S1 and S2**.

Focusing on geographic coverage to hospitals (**Table 2**); 71.6% of WoCBA lived within 30 minutes of a hospital offering childbirth care, considering average speed (63.9% for public hospitals). However, in the slowest speed scenario, this reduced to 22.9% for all hospitals and 18.9% for public hospitals. There were notable differences between health zones. For example, for a threshold of 30 minutes, the coverage ranged between 4.8% in Abomey-Calavi-Sô-Ava to 82% in Cotonou 1-Cotonou 4 for public hospitals and minimum speed scenario (**Table 2**). The percentage of WoCBA within 30-, 60- and 120-minute travel time thresholds for hospitals are shown by commune and *arrondissement* in **Supplementary Tables S1 and S2**.

**Table 2.**
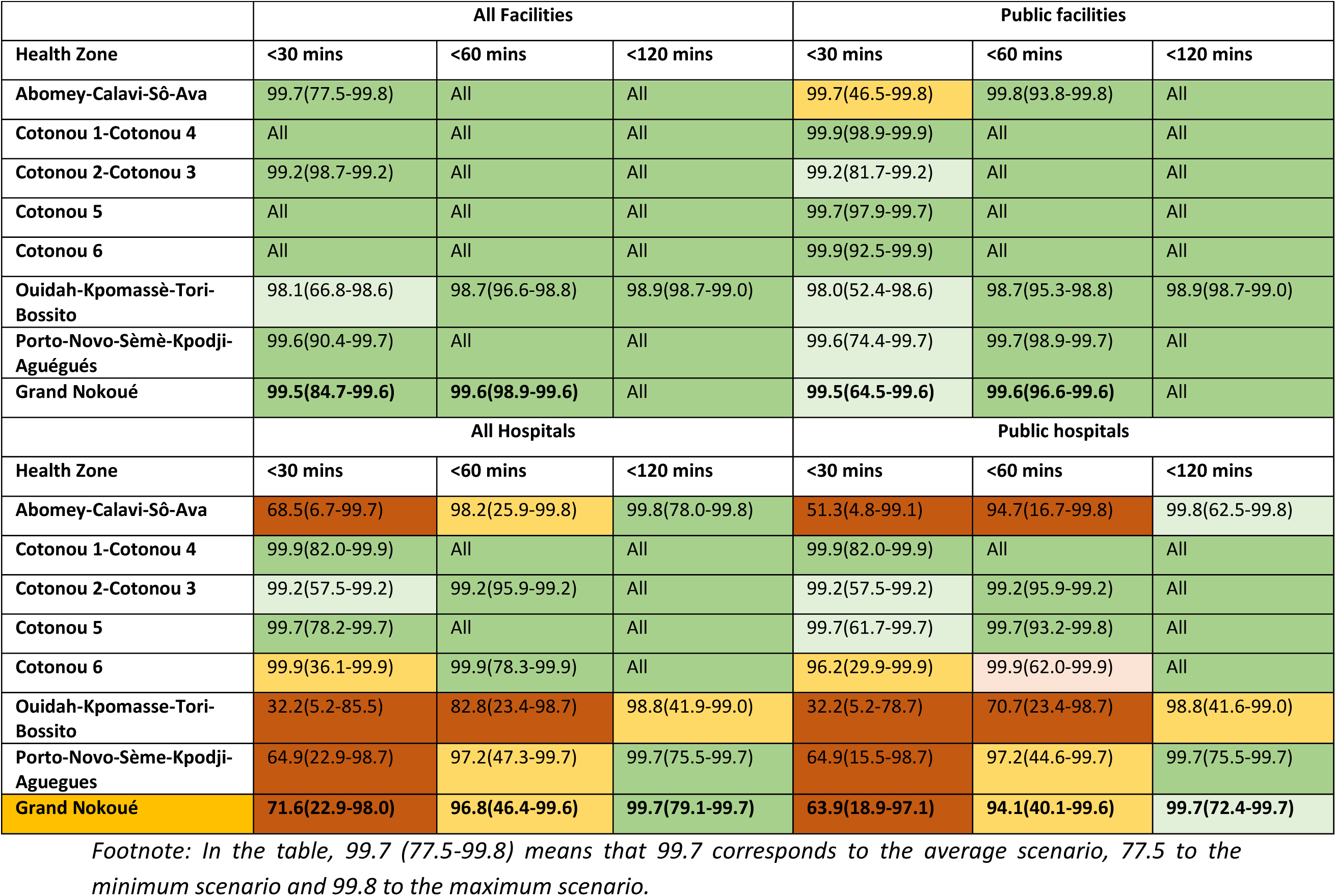
Percentage of women of childbearing age within 30, 60 and 120 minutes travel time to the nearest facility providing childbirth care, modeled by the three speed scenarios (average, maximum, minimum speed), by health zone.

### Wealth-related inequalities

Large pro-rich inequalities were observed in geographic access to hospitals, but were less pronounced for access to all facilities (**Fig 5)**. Specifically focusing on access to hospitals (all and public sector only) and looking at *Grand Nokoué* metropolitan area as a whole, women from the poorest quintile had the longest travel times to hospitals (50 minutes) compared to the richest quintile (10 minutes) in the average speed scenario. This pattern was also observed in Atlantique and *Ouémé* d*épartements*. However, in Littoral *département* the difference in travel time between different wealth quintiles groups was small and the middle quintile households had the best access (about 10 minutes to any hospital) followed by the richer and the richest while the poorer have the worst accessibility. The equiplots for other travel scenarios are presented in **Supplementary materials Figures S10 and S11.**

**Fig. 5.**
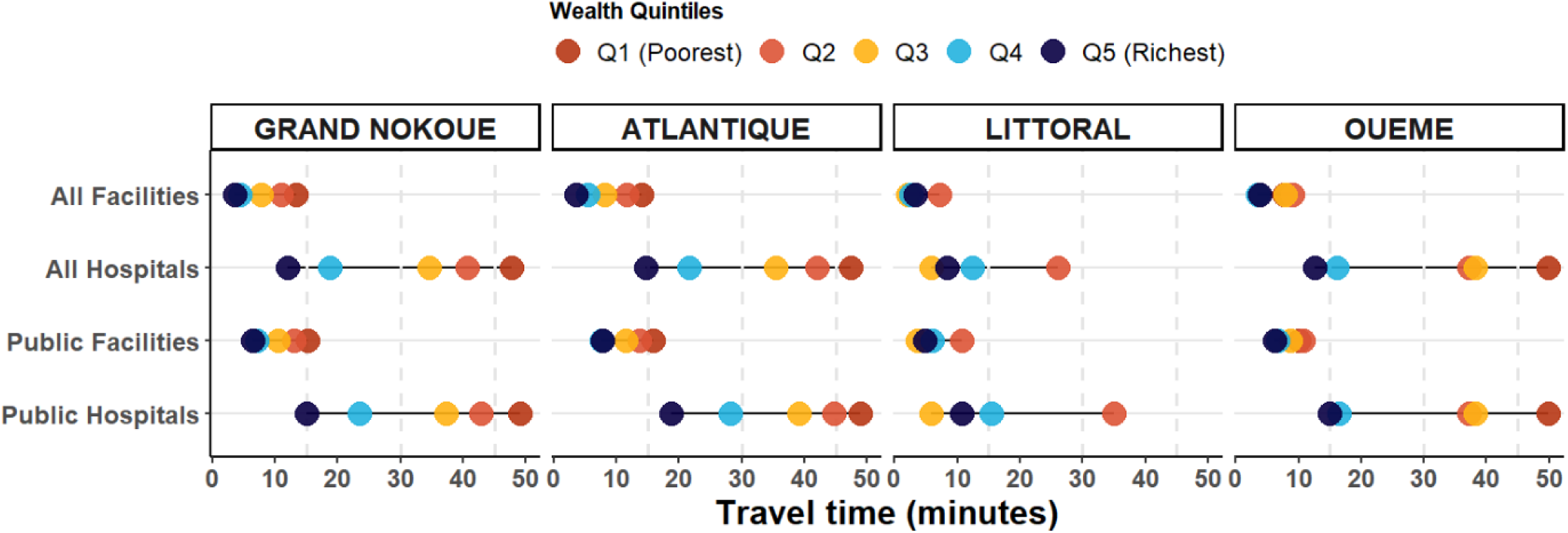
Equiplot of variation of travel time using average speed scenario to childbirth care relative to wealth index disaggregated by level of facility and *Département* within Grand Nokoué metropolitan area, Benin

### Urbanization gradient

The results of intersecting travel time and gridded surface of urbanization are shown in **Table 3**. Overall, irrespective of facility level or sector, the large and medium settlements had shorter travel times compared to peripheral areas containing small settlements, low density and almost uninhabited areas. For example, a hospital could be accessed within an average of half an hour in a city (core densely populated area), but this increased to almost 1 hour (52 minutes) in a village (small settlements) and 46 minutes in a very low-density areas (**Table 3).**

**Table 3.** Travel time across to health facilities offering childbirth services for different settlement types in the urbanicity continuum within *Grand Nokoué*, Benin.

| Settlements | All facilities | Public facilities | All hospitals | Public hospitals |
| --- | --- | --- | --- | --- |
| Travel speeds | Ave [max-min] | Ave [max-min] | Ave [max-min] | Ave [max-min] |
| City (large settlement) | 5.7 [2.7-19.3] | 8.6 [4.0-28.9] | 26.5 [12.0-89.3] | 30.2 [13.3-100.8] |
| Town (dense, medium settlement) | 5.0 [2.5-15.4] | 7.2 [3.4-22.9] | 35.0 [17-104.8] | 35.0 [17.0-104.8] |
| Town (semi dense, medium settlement) | 11.2 [5.7-33.1] | 11.2 [5.7-33.1] | 44.2 [20.7-120.5] | 44.2 [20.7-120.5] |
| Suburban or peri-urban (semi dense) | 13.1 [7.1-34.7] | 14.3 [7.6-38.9] | 39.6 [19.2-115.8] | 39.8 [19.3-116.5] |
| Village (small settlement) | 16.3 [9.1-41.6] | 16.4 [9.1-42] | 51.9 [25.9-137.9] | 52.0 [25.9-137.9] |
| Dispersed rural (low density) | 15.9 [8.8-40.4] | 16.7 [9.2-43] | 44.1 [21.8-126.1] | 44.5 [22.0-127.4] |
| Very low density (mostly uninhabited) | 19.2 [11-46.2] | 19.9 [11.3-48.6] | 46.2 [23.5-125.5] | 46.5 [23.7-126.2] |

## Discussion

We assessed geographical accessibility to facility-based childbirth care in *Grand Nokoué* metropolitan area using localised and context-specific travel speeds. We explored inequalities by disaggregating travel time estimates by wealth quintiles and urbanicity. Travel time to the nearest childbirth care facility in *Grand Nokoué* was relatively short (8 minutes) under average travel conditions. This increased substantially to 106 minutes for women needing public hospital care traveling at slowest speeds. Women from poorer households had five times longer travel times compared to their wealthier counterparts. Our work builds on and improves upon previous assessments of spatial access in Benin [4,5,20,22–25]. We contextualise these findings and their value to health system planning by identifying where patterns may inform evidence in *Grand Nokoué*.

Travel time to the most advanced and affordable care provided in public hospitals *-* 33 minutes in the average scenario, 106 minutes in the slowest speed scenario - was within the global threshold of 2-hours [6]. However, these durations still poses a risk as delays of 10 to 30 minutes have been associated with stillbirths and maternal mortality [7]. Overall, the travel time estimates in *Grand Nokoué* was slightly longer compared to other SSA cities where similar analyses were conducted [27,28]. Slow speeds and consequently long travel times in *Grand Nokoué* could be due to traffic congestion leading to long delays for women needing to access CEmONC. The actual time to accessing care for many women would be even longer, accounting for time to find transport, securing fees, reaching from the entrance of a health facility to the maternity ward, and additional time if referral to another facility is required.

Within *Grand Nokoué* travel time varied at *arrondissement* level, with eleven *arrondissements* where travel time was longer than 120 minutes at slowest speeds. Travel time was substantially shorter in Cotonou and Porto-Novo, two cities which are part of large settlements in the urbanization continuum, due to the high density of road networks and health facilities (70% of hospitals in *Grand Nokoué* are located in these two cities). On the other hand, Ouidah and its immediate neighbourhood (*Djègbadji, Houakpe-Daho, Avélékété*, *Pahou* and *Bossito*) had relatively longer travel times as the area has fewer facilities and a lower density of road network and some parts have water, flooded vegetation and trees, which limits movement.

Eight out of ten women in *Grand Nokoué* were within 30 minutes of any facility offering childbirth services at slowest speed. However, this declined to two out of ten within a half hour of a (public) hospital under the minimum speed, making geographic accessibility to hospital care, especially in critical situations, a significant barrier. *Arrondissements* like *Avelekete, Hèvié, Pahou, Ouèdo, Bossito*, had a geographic coverage of less than 15% of WoCBA within 2 hours. These *arrondissements* would benefit from targeted policy interventions to reduce women’s geographic marginalization from hospital-based childbirth care. Actions such as upgrading lower-level facilities to hospitals, improving of roads or providing alternative means of transport such as ambulances [20] would enhance the geographical coverage in the most affected *arrondissements*. In this line, the *Grand Nokoué* sustainable urban mobility project aims to improve urban mobility, accessibility, and safety along selected corridors and urban areas of the *Grand Nokoué*. This includes improving transport infrastructure (Abomey-Calavi – Cotonou and Ouidah – *Sèmè-Podji*), providing efficient and affordable bus-based public transport services along the main urban mobility corridor [29].

Our findings support the literature on the impacts of socio-economic status on access to childbirth care in SSA, whereby pro-rich inequalities have been documented in major cities [27,34]. The health inequalities in the country are exacerbated by socio-economic factors [19,21], considering that two-fifths of Beninese health expenditures are out-of-pocket [35]. Within *Grand Nokoué*, women from the poorest household wealth quintile travelled on average 5 times longer than those in the richest quintile to the nearest hospital. These pro-rich inequalities are due to a high distribution of hospitals in Cotonou and Porto-Novo, which also have households of relatively higher wealth. Almost half of the hospitals in *Grand Nokoué* are in Cotonou as a result of governmental efforts to improve access to healthcare. However, the population of Cotonou is reducing over time, as populations move toward other areas such as Abomey-Calavi and Sèmè-Kpodji. These peri-urban areas have a lower density of hospitals and therefore, approaches to decentralize, restructure or build new hospitals in these areas should be explored.

Although the private sector can be more expensive, it constitutes a vital alternative when public infrastructure is under pressure or takes too long to access. The private sector provides 45% of healthcare services in Benin [35], and up to 71% of childbirth services are private providers in *Grand Nokoué*. Neglecting the private sector in estimates of accessibility to care can underestimate the actual healthcare options available to the population and our analysis shows that travel time reduced significantly when comparing access to public facilities versus all (public and private) facilities.

Our findings highlight substantial disparities in geographical access to childbirth services according to level of urbanization as previously reported in Benin [23]. Travel time is shorter in large and medium settlements than peripheral areas containing small settlements, low density and almost uninhabited areas. The large and medium settlements areas are characterized by high population densities, well-maintained roads, efficient public transportation, and proximity to vital services. Conversely, peripheral areas have poor transportation infrastructure and fewer public transportation options.

The spatial metrics of travel time generated in this study relied heavily on travel speeds across different road types. Some road types saw a diverse traffic mix, including cars, cyclists and pedestrians, requiring slower speed limits for safety reasons. Most of these were interurban roads where people live, such as in Abomey Calavi-Allada. Additionally, during the data collection period, road construction and resurfacing work was underway in *Grand Nokoué*, which led to diverting road users and reducing the speed of vehicles om in some areas. The speeds recorded in *Grand Nokoué* were similar to those recorded in Grand Conakry, Guinea [27] but are much slower than the generic speeds where major roads are assigned 100 km/hr [4,5]. Therefore, it is likely that previous studies have overestimated geographic accessibility especially in urban settings when generic speeds have been used.

### Strengths

We used locally measured speeds reflecting the ground reality in *Grand Nokoué.* The approach used to collect travel speeds is inexpensive, scalable and can easily be embedded in ongoing research activities. Further, we provided a continuum of uncertainty from the slowest to the fastest speed unlike the majority of spatial accessibility analyses that only provide a point estimate. Second, we included the entire *Grand Nokoué* metropolitan area with various degrees of urbanization. This allowed inclusion of women living on the periphery who would gravitate toward Cotonou for healthcare as seen in other metropolitan areas in SSA such as Kampala and Conakry [27]. Third, we included facilities from public and private sectors and we validated those that offer childbirth services with routine data unlike analysis that incorporate the public sector only. Fourth, we disaggregated travel time estimations across subnational units used for decision making within the metropolitan area and also explored how travel time varied by wealth and urbanization to improve our understanding of the inequalities.

### Limitations

First, we computed travel time to the nearest facility, however, we acknowledge that women may bypass nearest facilities, even in an emergency, and may have led to overestimating accessibility. Second, to improve understanding of inequalities within the metropolis, we disaggregated travel time estimates based on relative wealth index and urbanicity gradient. However, we did not disaggregate based on informal settlements due to lack of the corresponding geospatial data despite that 64% of urban Beninese population live in slums or informal settlements. Third, it is also possible that some women within *Grand-Nokoué* seek care outside *Grand-Nokoué* due to easier access to health facilities in the peripheral areas. Fourth, travel speeds were geotraced during the dry season, therefore it’s likely that travel speeds would be much lower in the rainy season leading to longer travel times. Fifth, our estimates are based OSM road network with a completeness of 82%. However, roads in peri-urban areas are not fully digitally captured in volunteered geographic information which may have led to biased (longer) travel time in the periphery. Finally, we did not consider the quality of the roads and adverse weather conditions such as extreme rainfall which could have led to longer travel times.

## Conclusions

Within the *Grand Nokoué* metropolitan area, average geographic accessibility to and coverage for facilities offering childbirth care is adequate. However, at slowest speeds, some areas and populations have travel times of over two hours. Pro-rich inequalities are widespread, especially to hospital childbirth care. These findings are relevant to policy makers for spatial targeting and prioritization of those with a dual burden (poorest wealth and living far away) such as those in peri-urban areas and suburbs. Additionally, and well-maintained roads, regulating transportation to reduce congestion, building hospitals and/or upgrading facilities and providing ambulances could be considered by relevant stakeholders.

## Supporting information

Grand Nokoue supplementary file

## Data Availability

All data produced in the present study are available upon reasonable request to the authors

## Acknowledgements

We are indebted to the support of researchers from the *Centre de recherche en reproduction humaine et en démographie* (Maxime Houndodjade, Thierry Dannon; Anissa Abdoulaye, Harziki Boukari) who volunteered and supported Yiséché Hounménou and Elias Avahoundjè, in geotracing travel speeds within the *Grand Nokoué* metropolitan area.

## Ethical approval

This study used secondary publicly available data and primary data (Travel speeds, location of health facilities) which did not require ethical approval.

## Funding

The study was funded by Fonds voor Wetenschappelijk Onderzoek (FWO) Grant ID: G074724N and The Belgian Federal Directorate-General for Development Cooperation Humanitarian Aid (DGD). PMM is supported by FWO for his Senior Postdoctoral Fellowship (#1201925N). The funders had no role in study design, data collection and analysis, decision to publish, or preparation of the manuscript

## Data

The datasets used for this analysis are publicly available. These include a database of health facilities obtained from the Ministry of Health and Public Hygiene, while links for population relative wealth index and factors affecting travel to healthcare (road network, digital elevation model and land use) are listed in the manuscript.

## Competing interests

The authors declare they have no competing interests.

## Authors’ contribution

[utable]

## Notes

### Competing Interest Statement

The authors have declared no competing interest.

### Funding Statement

This study was funded by Fonds voor Wetenschappelijk Onderzoek (FWO) Grant ID: G074724N and The Belgian Federal Directorate-General for Development Cooperation Humanitarian Aid (DGD). PMM is supported by FWO for his Senior Postdoctoral Fellowship (#1201925N).

