## Supplementary material for "Modeling geographical accessibility and inequalities to childbirth services in the *Grand Nokoué* metropolitan area, Benin": Grand Nokoue supplementary file

#### Section 1: Benin and Grand Nokoué context

##### Healthcare service delivery

The health service delivery platform in Benin is pluralist (with public and private health care and service providers) and has a pyramidal structure. Health facilities which provide childbirth care include, in order from lowest to highest level, a network of first-contact public services (maternity and health centers) and private health facilities, supported by a first-referral public or private hospital namely health zone hospitals (*Hôpital de zone* / *Centre hospitalier universitaire de zone* - CHUZ); departmental hospitals (*Centre hospitalier départemental* - CHD/ *Centre hospitalier universitaire départemental*- CHUD); and the national hospital (*Centre national hospitalier et universitaire Hubert Koutoukou Maga* – CNHU-HKM/ *Centre hospitalier universitaire de la mère et de l'enfant lagune*- CHU-MEL) within *Grand Nokoué*. In addition, the private health sector in Benin exists in two forms: for-profit and not-for-profit. The for-profit sector includes medical and paramedical practices, clinics, polyclinics and private diagnostic centers or medical centers. The not-for-profit sector includes mainly confessional health centers or FBOs health facilities and NGO facilities. Most of health centers offer childbirth and sometimes neonatal healthcare. In Cotonou (the core urban area), almost all women attend at least one antenatal care visit (98.6%) and give birth in a health facility (99.2%) [1]. Among the women give birth in facility, 53.2% are in the public sector and 46.0% in the private sector [1,2].

Within the conurbation, the existing road network totals approximately 5,500 km in 2023, of which 20% (1,100 km) are tarmacked [3]. Walking is the primary means of transportation [3], however, the conurbation is witnessing a rise in traffic congestion due to the inadequacy of urban road infrastructure and traffic management system [3]. Motorbikes account for approximately 70% of motorized journeys, which also contributes to the congestion [3]. Women face a combination of challenges when accessing childbirth care such as high cost of transport and delays caused by traffic jams poor quality of roads and uncomfortable transport options such as use of a motorbike for WoCBA while pregnant, in labor, or with a complication [4]. These challenges are further exacerbated by extreme climatic events, such as recurrent flooding, especially during the rainy season [5].

##### Subnational boundaries

The *Grand Nokoué* covers an area of about 1,444 $km^2$, approximately 1.2% of Benin's total area (114,763 $km^2$). Despite its relatively small size, *Grand Nokoué* is densely populated, hosting about 2.38 million inhabitants in 2020 [3] (about 20% of Benin's population and 80% of the urban population). This population is projected to reach over 3 million by 2030 reflecting rapid demographic growth in the region [3].

The *Grand Nokoué* consists of five administrative communes: Cotonou (Benin's economic capital), Porto-Novo (Benin's administrative capital), Ouidah (tourist center), Abomey-Calavi (Benin's industrial hub), and *Sèmè-kpodji* (business and industrial center). Each commune is subdivided into several *arrondissements*, making a total of 43 *arrondissements* in the *Grand Nokoué area*. In terms of health service delivery, the *Grand Nokoué* consists of seven health zones (**Figure S1**), which are part of three administrative regions (**Figure S2**) known as (*Département* of Atlantique, Littoral, and *Ouémé*).

**Figure S1:** A map of 7 health zones within the *Grand Nokoué* metropolitan area in Benin

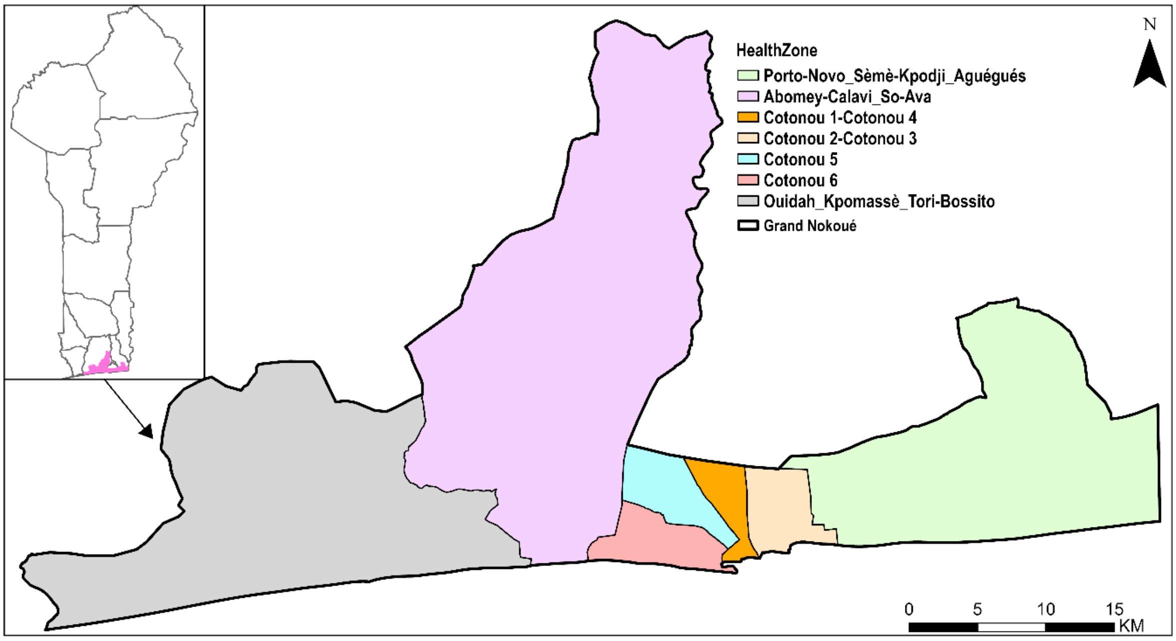


**Figure S2:** A map of 3 departments within the *Grand Nokoué* metropolitan area in Benin

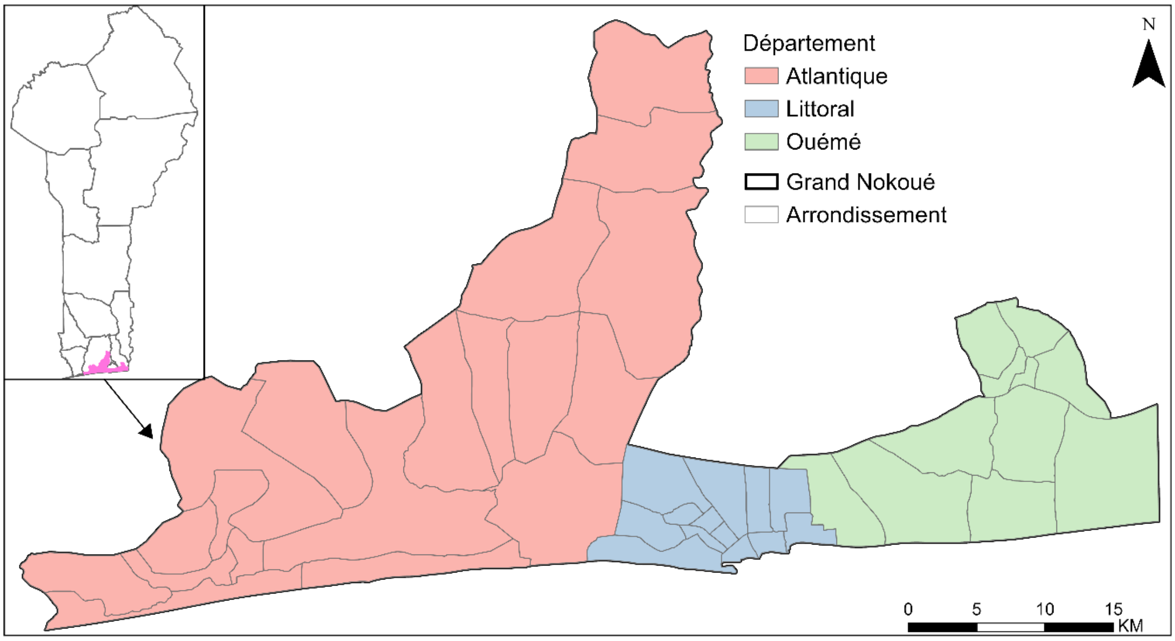

#### Section 2: Data

##### *Health facilities*

We required a geocoded database of health facilities that offer childbirth care in *Grand Nokoué* in 2023 (most recent). We achieved this by intersecting two databases of health facilities. The first database was obtained from Benin's health information system through the DHIS2 (formerly district health information system version 2). We extracted records of health facilities where at least one case of childbirth had been recorded between 2020 and 2023, including the level and the sector of the facility. To add a location attribute (latitude and longitude) to the assembled list, we relied on a database of health facilities from the Ministry of Health (Administration and Finance Programming Department, DPAF) and merged the two databases based on facility names and their corresponding administrative units. Where coordinates were not available, we geolocated based on online gazetteers including Google Maps, OpenStreetMap (OSM), and GeoNames. In this study, we combined facilities managed by the government and FBOs, hereafter referred to as public facilities, because their non-profit nature and financial accessibility (user fees) is similar while the for-profit are henceforth referred to as private facilities (hospitals, clinics and medical centres).

##### *Road network:*

The road network provides the transportation infrastructure needed to travel between residential areas and facilities providing childbirth care. We downloaded the most recent version (2024) of the road network in Benin from OSM [6]. We retained the road network corresponding to the spatial extent of *Grand Nokoué*. We adopted the hierarchy of road classes (trunk, primary, secondary, tertiary, residential, service, unclassified, and others) as defined by OSM and based on sections of roads which were geo-traced as described in the travel speeds section. The road network is shown in **Supplementary figure S3**

**Figure S3:** A map of the road network within the *Grand Nokoué* metropolitan area in Benin

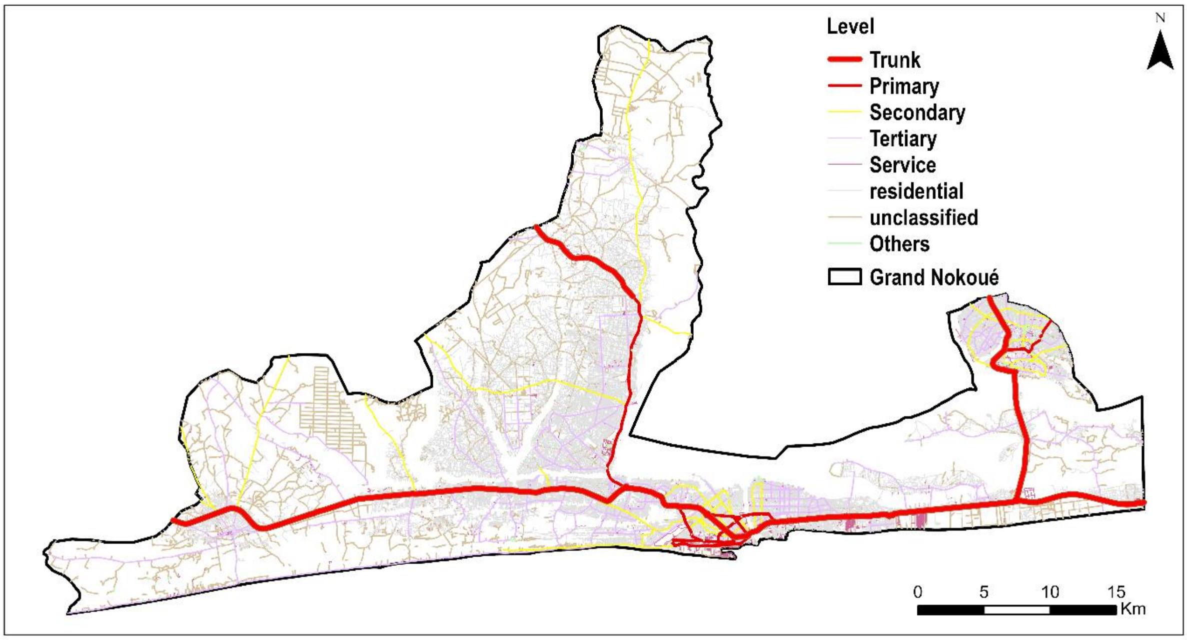

##### ***Land use/cover:***

The land use/cover was used to represent areas between roads. The land use/cover is derived from European Space Agency Sentinel-2 imagery based on a deep learning model trained by human-labeled image pixels with an overall accuracy of over 91%  [7,8]. The land use/cover map was available at 10 × 10 m spatial resolution for 2023 from the ArcGIS living atlas of the world [9]. The data contained seven land use/cover classes: water, trees, flooded vegetation, crops, built-up areas, and rangeland (**Figure S4**). Flooded vegetation and water were considered as transport barriers, except where there was a bridge.

**Figure S4:** A map of the land use/cover within the Grand Nokoué metropolitan area in Benin

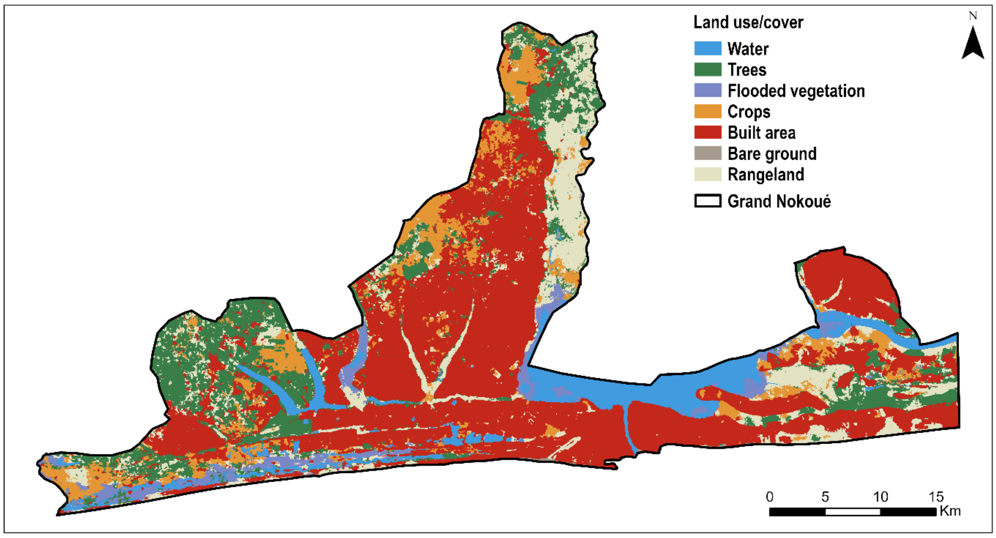


##### **Digital elevation model (DEM):**

Walking speeds vary by slope of the terrain. The Shuttle Radar Topographic Mission DEM (**Figure S5**) at the 30 × 30 m spatial resolution [10] was to derive the slope. The slop in turn was used to modify the walking speeds based on Tobler's formulation, an exponential function that describes how human walking speed varies with slope [41].

**Figure S5:** Digital elevation model  of the *Grand Nokoué* metropolitan area in Benin

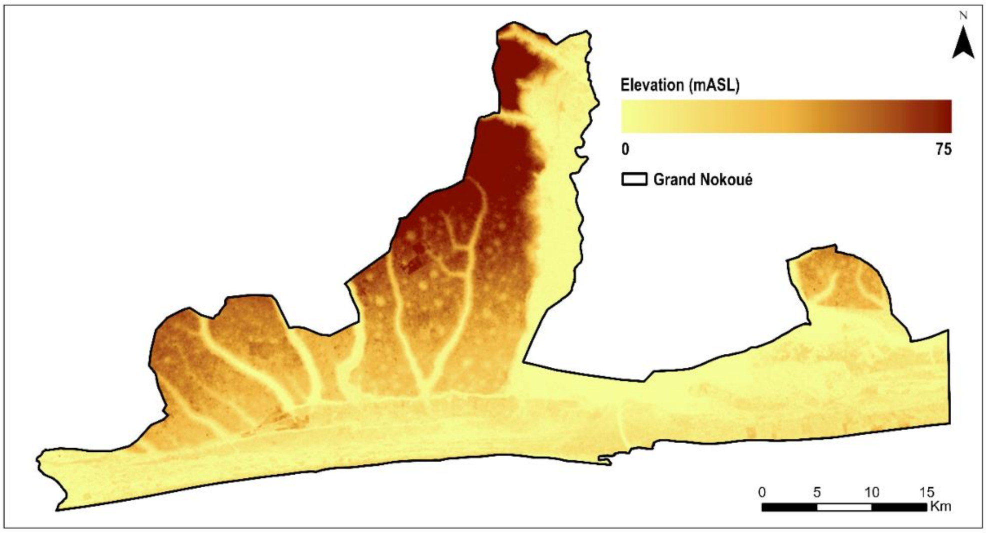

**Relative wealth index (RWI):**

The RWI is a proxy for relative standard of living constructed using machine learning approaches for 93 low and middle-income countries [12]. It is based on de-identified connectivity data (such as cell towers, Wi-Fi access points, and mobile devices from Meta), satellite imagery, nightlights, population, elevation, slope, built up areas, and road density. The data is provided at 2.4 km spatial resolution, and validated based on a variety of datasets such as household surveys, census and Gross Domestic Product [12]. The data was available from the Humanitarian Data Exchange portal [13] and shown in **Supplementary figure S7.**

**Figure S6:** Relative wealth index within Grand Nokoué metropolitan area in Benin

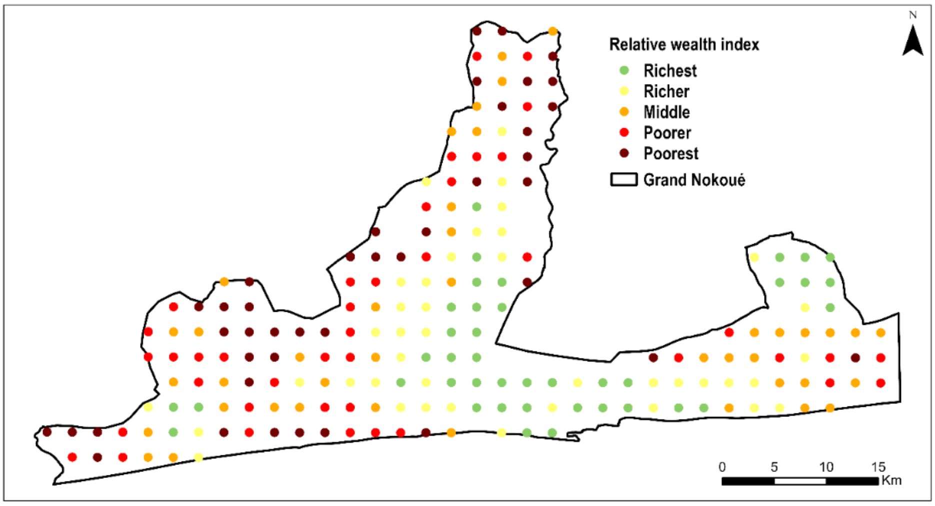


##### *Urbanicity gradient*

Settlements (in the continuum of a hamlet to a mega-city) across the globe have been classified according to the Degree of Urbanization concept [14] based on population size and density and built-up area densities. It classifies an area (1 x 1 km spatial resolution) into 7 classes: city (large settlement), town (either dense or semi-dense), suburban/peri-urban, villages (small settlement), dispersed rural area (low density area) and mostly uninhabited area (very low density area). The dataset was downloaded from Copernicus' Global Human Settlement Layer portal [15] and shown in **Figure S8.**

**Figure S7:** Urbanicity gradient within *Grand Nokoué* metropolitan area in Benin

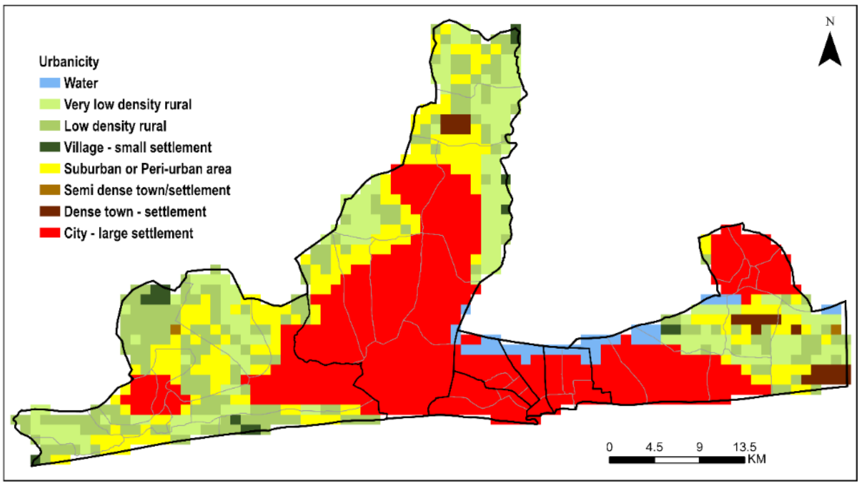

##### ***Population of women of childbearing age (WoCBA)***

We derived estimates of the number of WoCBA in the study area based on high-resolution gridded datasets available from Worldpop's open spatial demographic data and research portal [16]. Specifically, we downloaded estimates of number of people per 100m grid square stratified by sex and 5-year age groups for 2020, constrained within human settlements [17]. This version adjusts country totals to match the corresponding official United Nations population estimates. We summed the counts for females aged 15 to 49 years to derive a 2020 map of WoCBA at 100m spatial resolution. We then obtained the 2023 projected counts of WoCBA at the commune level within *Grand Nokoué* provided in the United States Census Bureau portal for Benin [18]. We spatially matched these totals at the commune level with the gridded surface of 2020 while maintaining the 100m spatial resolution resulting in a 2023 projected surface of WoCBA in *Grand Nokoué* **(Figure S8).**

**Figure S8:** Population distribution of women of child bearing age within *Grand Nokoué* metropolitan area in Benin

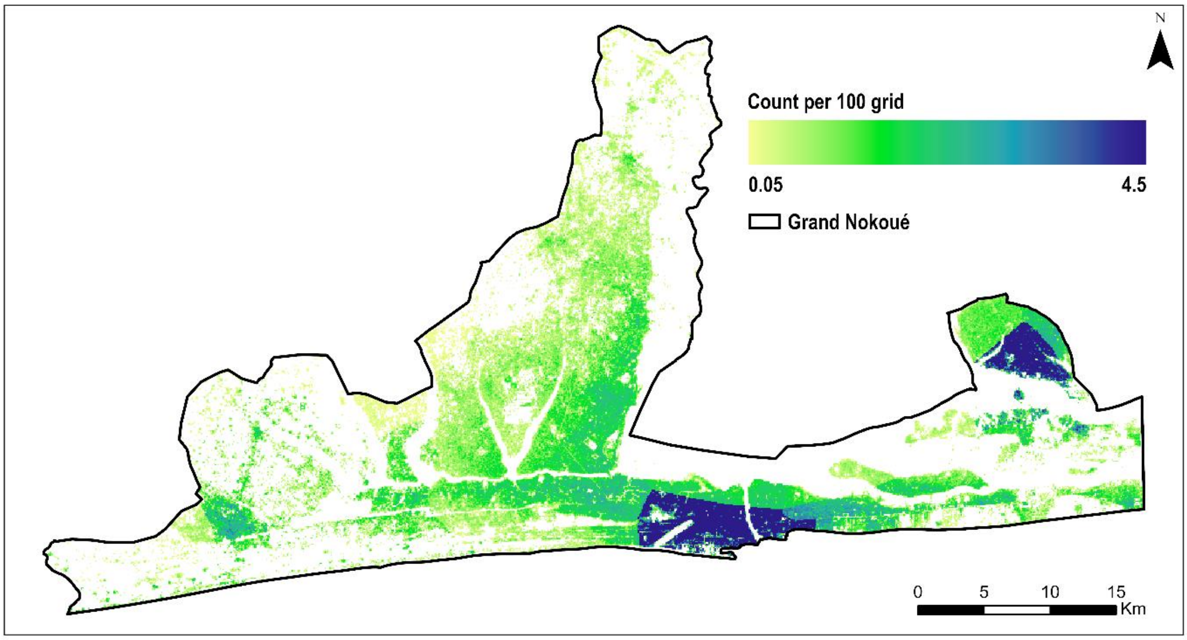


#### Section 3: Travel speeds

We used a previously validated approach to obtain motorized travel speeds for different road types in Grand Conakry, Guinea [19], to obtain the travel speeds in the *Grand Nokoué* metropolitan area. This approach provides local context speeds, more granular relative to generic speeds routinely used in analyses of accessibility [42].

Briefly, from June 14th to July 29th, 2024, seven volunteers working at the *Centre de recherche en reproduction humaine et en démographie,* Benin geo-traced their motorized journeys (trajectories) within the study area at different times of the day using tablets or smartphones with the KoboCollect App [43]. Within the app's *geotrace* option, global positioning system (GPS) points were taken along each trajectory, providing a location every minute with an accuracy of 20 meters. At the beginning and end of each trajectory, the data collector was prompted to enter start and end time.

For each trajectory, we collected additional attributes, including mode of transportation, road type and its quality. Data were downloaded from the KoboCollect App as a spreadsheet. Overall, 110 trajectories were completed. The trajectories were validated in the following three steps. First, we removed four trajectories without GPS coordinates. Second, for the remaining trajectories, we calculated the travel time taken by the data collector per trajectory in two ways. As the difference between start and end time (T1) and the time obtained by considering the total number of geo-traced points every minute by the application (T2). Ideally T1 and T2 should be equal; that is the number of geo-traced points (every minute) should be equal to the time taken between the first and geo-traced points for a single trajectory. If the difference between T2 and T1 was less than 3 minutes, the trajectory was considered valid. If the difference was between 4 and 10 minutes and T2 was greater than four times the difference, the route was also valid. All the journeys that did not meet this condition (n=56) were considered to be non-valid and placed at the third level of validation. At the third level, we mapped and displayed the 56 routes using ArcGIS Pro v 3.3.1 (ESRI, Redlands, CA, USA). Based on our local knowledge of the setting, we checked closely whether there were any inconsistencies linked to each of them leading to exclusion of 31 routes. In the end, 75 routes were retained for inclusion in the analysis.

We then computed the average travel speed on each road segment as the ratio between total distance and the time taken across the segment (T2). Finally, we linked the estimated speeds with the corresponding road segment on the OSM road network by spatially overlaying the traced GPS markers with the road network. The combination of OSM road segment type and speeds was summarized using mean (moderate case scenario, average travel disruptions), slowest (worst case scenario, high travel disruptions), and fastest (best case scenario, travel disruptions).

#### Section 4: Modelling travel time

We modelled travel time to the nearest facility offering childbirth care based on the least cost path algorithm. The least cost path was defined as the path that would take the least amount of time to reach a facility while accounting for the mode of transport, speed, road network, topography, and travel barriers. Travel time was estimated from 30m square girds to all facilities and separately, to all hospitals and a subset of facilities by level in the public sector. For each of four categories of health facilities, three models were estimated using different travel speeds; the slowest and fastest speed to provide a continuum of the worst and the best travel scenarios, as well as the average speed. The least-cost path algorithm was implemented in AccessMod software version 5.8.0, a WHO tool used to model spatial accessibility and coverage [22,23].

Specifically, we first merged the land use/cover and the road network using the *"merge land cover module"* in AccessMod. We then applied different travel speeds on the resultant merged surface to define travel time from each 30m square grid in *Grand Nokoué* to the nearest facility for the four defined subsets of facilities (disaggregated by level and by sector). We defined a hybrid mode of transport where walking or motorized transport can be used. In areas without a road network, a person will walk until where there is a road network for motorable transport. The walking speeds were variable depending on the type of land cover/use and modified based on slope [11], the so-called Anisotropic option in AccessMod. In implementing the least cost path, we specified the "Knight's move" that allows determination of the least cost path based on 16 neighboring cells instead of the default eight cells leading to more accurate results despite a higher computational expense.

We then masked non populated areas in the resultant gridded surface of travel time by use constrained population distribution maps. We used the zonal statistics function of ArcGIS Pro v 3.3.1 (ESRI, Redlands, CA, USA) to summarize the average travel time across the *Grand Nokoué* Metropolitan areas and sub-nationally for each health zone, commune and *arrondissement*.

### Results

**Figure S8:** Travel time to childbirth care in all facilities and public sector facilities at 30 m spatial resolution, by travel speed scenario (minimum, average and maximum) in *Grand Nokoué* metropolitan area, Benin

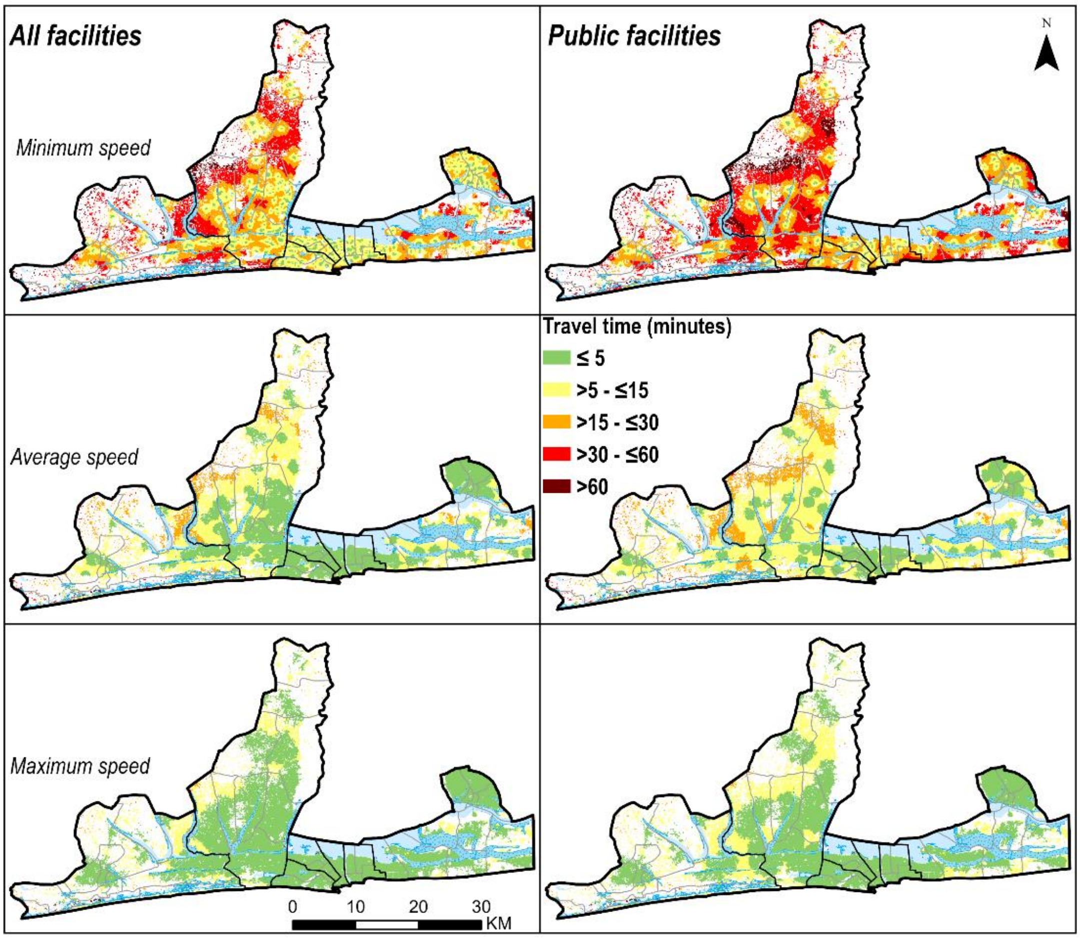

**Figure S9:** Travel time to childbirth care in all hospitals and public sector hospitals at 30 m spatial resolution, by travel speed scenario (minimum, average and maximum) in *Grand Nokoué* metropolitan area, Benin

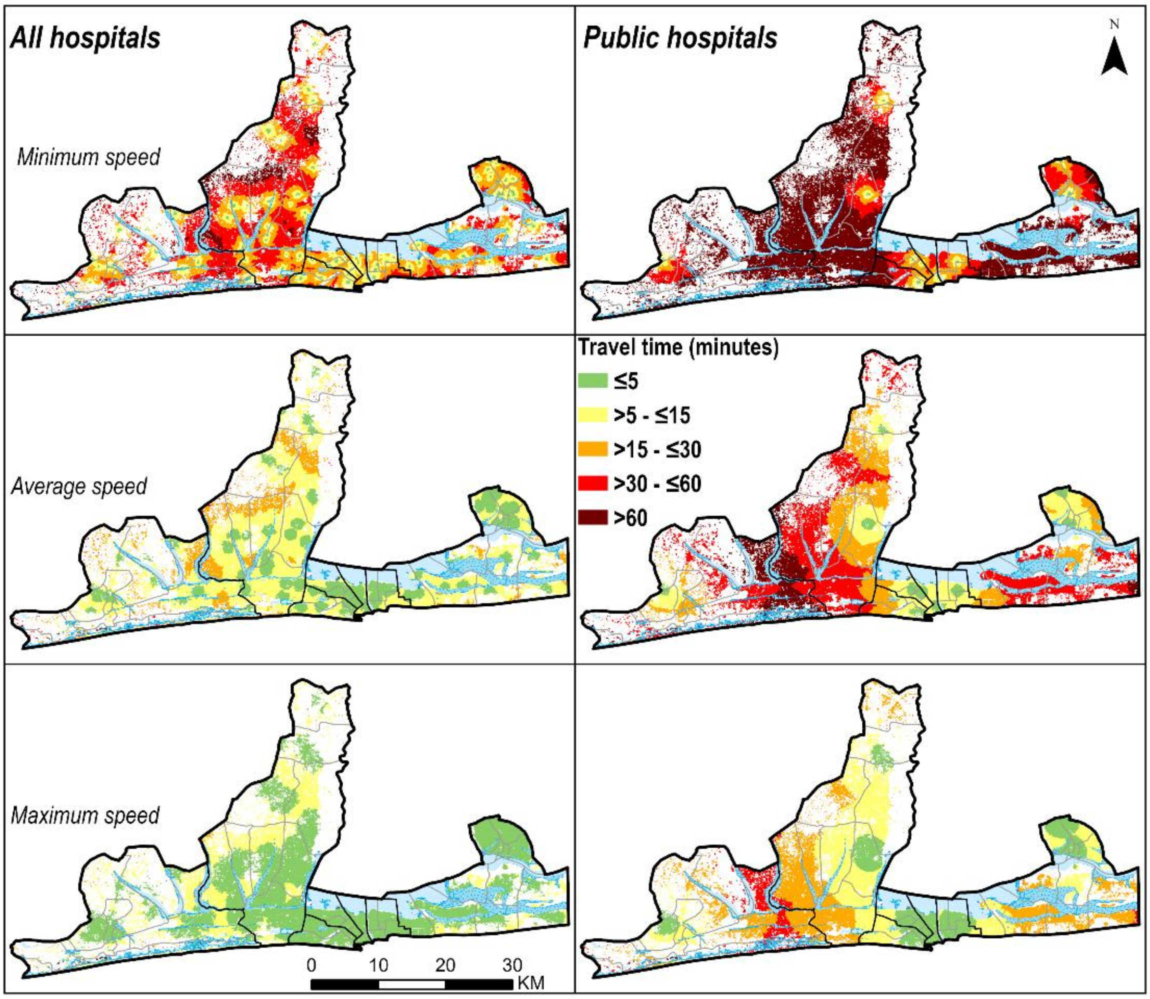


**Figure S10:** Equiplot of variation of travel time to childbirth services using maximum speed scenario relative to wealth index disaggregated by level of facility and departments within *Grand Nokoué*, Benin in 2023

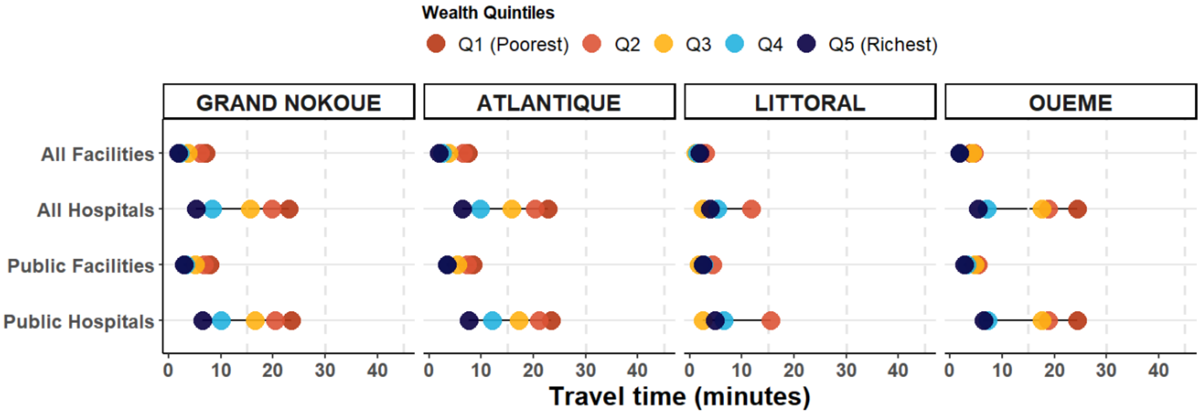

**Figure S11:** Equiplot of variation of travel time to childbirth services using minimum speed scenario relative to wealth index disaggregated by level of facility and departments within *Grand Nokoué*, Benin in 2023

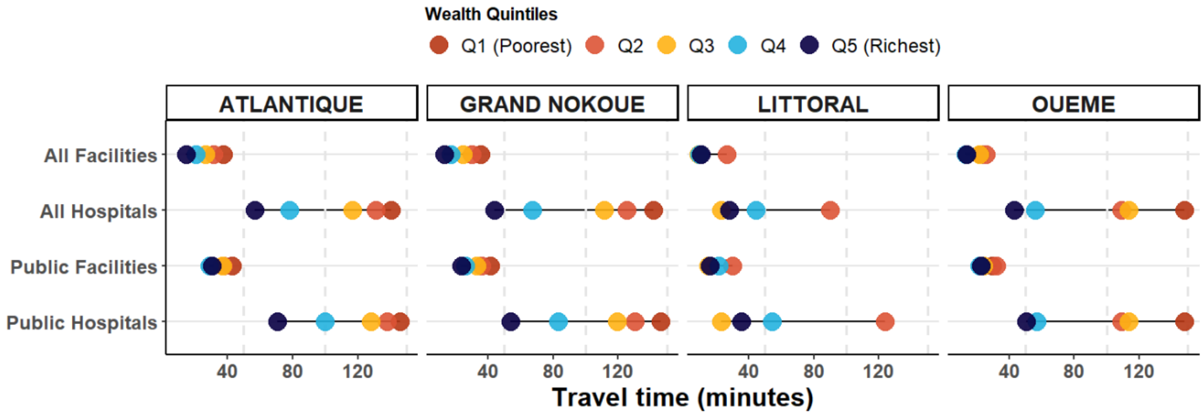

**Table S1**: Proportion of women of childbearing age within 30, 60 and 120 minutes of travel time to all facilities and hospitals by a*rrondissement*

| | | All Facilities | | | All Hospitals | | |
|---|---|---|---|---|---|---|---|
| **Arrondissement** | **Population** | **<30 mins** | **<60 mins** | **<120 mins** | **<30 mins** | **<60 mins** | **<120 mins** |
| Akassato | 43976 | 99.7(42.4-99.8) | 99.8(98.8-99.8) | All | 90.0(7.5-99.8) | 99.8(28.6-99.8) | 99.8(98.4-99.8) |
| Calavi | 67567 | 99.9(91.3-100.0) | All | All | 99.9(20.2-99.9) | 100.0(57.2-100.0) | All |
| Glo Djibe | 18534 | 99.9(62.1-99.9) | 99.9(98.8-99.9) | All | 38.1(0.0-99.9) | 99.8(2.2-99.9) | 99.9(75.4-99.9) |
| Godomey | 100747 | 99.6(94.9-99.6) | All | All | 76.4(1.6-99.6) | 99.6(23.2-99.6) | 99.6(89.0-99.6) |
| Hèvié | 35721 | 99.7(63.0-99.9) | 99.9(97.3-99.9) | All | 0.0(0.0-98.8) | 85.2(0.0-99.9) | 99.9(0.0-99.9) |
| Kpanroun | 3754 | 97.9(40.4-99.7) | 99.7(98.0-99.7) | All | 18.8(0.0-97.9) | 99.5(0.0-99.7) | 99.7(62.8-99.7) |
| Ouèdo | 14992 | 100.0(66.3-100.0) | 100.0(94.6-100.0) | All | 0.0(0.0-100.0) | 100.0(0.0-100.0) | 100.0(1.5-100.0) |
| Togba | 34267 | 100.0(75.5-100.0) | All | All | 71.0(0.0-100.0) | 100.0(9.3-100.0) | 100.0(88.7-100.0) |
| Zinvie | 8773 | 98.6(80.8-99.6) | 99.6(98.5-99.6) | All | 98.5(38.3-99.6) | 99.6(78.3-99.6) | All |
| **ABOMEY-CALAVI** | **328331** | **99.7(77.5-99.8)** | **All** | **All** | **68.5(6.7-99.7)** | **98.2(25.9-99.8)** | **99.8(78.0-99.8)** |
| All 1er arr. | 17231 | 99.9(98.4-99.9) | All | All | 99.9(32.1-99.9) | 99.9(90.2-99.9) | All |
| 2eme arr. | 10016 | All | All | All | 99.9(96.5-99.9) | 99.9(99.9-99.9) | All |
| 3eme arr. | 11849 | 98.2(98.2-98.2) | 98.2(98.2-98.2) | 98.2(98.2-98.2) | 98.2(39.9-98.2) | 98.2(98.2-98.2) | 98.2(98.2-98.2) |
| 4eme arr. | 11309 | 98.5(98.4-98.5) | 98.5(98.5-98.5) | 98.5(98.5-98.5) | 98.5(80.2-98.5) | 98.5(98.5-98.5) | 98.5(98.5-98.5) |
| 5eme arr. | 11354 | All | All | All | All | All | All |
| 6eme arr. | 14074 | All | All | All | 99.9(67.5-99.9) | All | All |
| 7eme arr. | 9762 | All | All | All | 100.0(82.9-100.0) | All | All |
| 8eme arr. | 10669 | All | All | All | 100.0(97.2-100.0) | All | All |
| 9eme arr. | 15257 | All | All | All | 99.1(70.5-99.2) | All | All |
| 10eme arr. | 11404 | All | All | All | 100.0(66.6-100.0) | All | All |
| 11eme arr. | 9784 | All | All | All | 100.0(73.1-100.0) | All | All |
| 12eme arr. | 30348 | All | All | All | 99.8(38.9-99.8) | 99.8(61.0-99.8) | All |
| 13eme arr. | 23289 | All | All | All | 100.0(17.0-100.0) | 100.0(91.7-100.0) | All |
| **COTONOU** | **186346** | **All** | **All** | **99.7(58.8-99.7)** | **99.7(91.4-99.7)** | **All** | **All** |
| 1 er arr. | 1392 | 99.8(27.0-100.0) | All | All | 94.0(0.0-100.0) | 100.0(0.0-100.0) | All |
| 2 eme arr. | 10028 | 100.0(93.3-100.0) | All | All | 99.8(32.4-100.0) | 100.0(96.1-100.0) | All |
| 3 eme arr. | 6772 | 100.0(67.4-100.0) | All | All | 99.4(0.0-100.0) | 100.0(86.9-100.0) | All |
| 4 eme arr. | 3091 | All | All | All | 99.3(0.0-99.7) | 99.7(29.0-99.7) | All |
| Avélékété | 2644 | 89.9(49.7-92.6) | 92.9(81.2-92.9) | 93.6(93.0-93.9) | 0.0(0.0-55.5) | 48.6(0.0-92.9) | 92.9(0.0-93.7) |
| Bossito | 3352 | 99.1(33.4-99.8) | 99.8(87.0-99.8) | All | 0.0(0.0-70.8) | 63.8(0.0-99.8) | 99.8(14.8-99.8) |
| Djegbadji | 2614 | 80.6(33.8-83.9) | 85.0(79.4-85.8) | 87.5(86.0-88.8) | 3.9(0.0-68.7) | 64.7(0.0-85.0) | 86.6(35.7-88.0) |
| Houakpe_ Daho | 989 | 62.2(40.5-80.3) | 82.6(54.6-84.9) | 88.2(78.2-89.6) | 0.0(0.0-63.4) | 62.1(0.0-83.3) | 86.4(51.7-89.0) |
| Pahou | 36918 | 99.4(67.3-99.5) | 99.5(98.9-99.5) | All | 0.4(0.0-78.8) | 74.6(0.0-99.5) | 99.5(2.3-99.5) |
| Savi | 9629 | 99.2(60.3-99.7) | 99.7(96.7-99.7) | All | 37.3(8.0-99.7) | 99.7(17.3-99.7) | 99.7(87.5-99.7) |
| **OUIDAH** | **77429** | **98.1(66.8-98.6)** | **98.7(96.6-98.8)** | **98.9(98.7-99.0)** | **32.2(5.2-85.5)** | **82.8(23.4-98.7)** | **98.8(41.9-99.0)** |
| 1e Arr. | 26461 | All | All | All | 99.6(11.6-99.9) | 99.9(77.9-99.9) | All |
| 2e Arr. | 6031 | 98.5(98.5-98.5) | 98.5(98.5-98.5) | 98.5(98.5-98.5) | 98.5(98.5-98.5) | 98.5(98.5-98.5) | 98.5(98.5-98.5) |
| 3e Arr. | 6595 | All | All | All | All | All | All |
| 4e Arr. | 22726 | All | All | All | 100.0(71.7-100.0) | All | All |
| 5e Arr. | 16114 | All | All | All | 100.0(51.2-100.0) | All | All |
| **PORTO-NOVO** | **77927** | **99.8(96.8-99.8)** | **All** | **All** | **99.7(51.5-99.8)** | **99.8(92.4-99.8)** | **All** |
| Aglangandan | 22190 | 99.2(93.7-99.2) | All | All | 94.7(0.7-99.2) | 99.2(25.7-99.3) | 99.3(97.7-99.3) |
| Aholouyeme | 3355 | 99.2(39.3-99.3) | All | All | 12.4(0.0-98.7) | 99.0(0.0-99.5) | 99.5(73.5-99.7) |
| Djeregbe | 13805 | 99.8(81.6-99.8) | All | All | 98.4(2.8-99.8) | 99.8(45.1-99.8) | All |
| EkpE | 28219 | 99.3(88.4-99.4) | All | All | 7.0(0.0-99.3) | 99.3(0.0-99.4) | 99.4(35.9-99.4) |
| SEmE - Kpodji | 12817 | 99.8(89.0-99.8) | All | All | 0.1(0.0-99.8) | 99.8(0.0-99.8) | 99.8(22.1-99.8) |
| Tohoue | 19055 | 99.6(79.5-99.7) | 99.7(93.5-99.7) | All | 1.8(0.0-91.1) | 77.1(0.0-99.7) | 99.7(27.5-99.7) |
| **SEME PODJI** | **99441** | **99.5(85.4-99.5)** | **99.5(98.3-99.5)** | **All** | **37.6(0.6-97.8)** | **95.2(12.0-99.5)** | **99.5(56.4-99.6)** |

**Table S2**: Proportion of women of childbearing age within 30, 60 and 120 minutes of travel time to all public facilities and public hospitals by a*rrondissement*

| | | Public facilities | | | Public hospitals | | |
|---|---|---|---|---|---|---|---|
| **Arrondissement** | **Population** | **<30 mins** | **<60 mins** | **<120 mins** | **<30 mins** | **<60 mins** | **<120 mins** |
| Akassato | 43976 | 99.7(29.3-99.8) | 99.8(88.4-99.8) | All | 79.6(3.2-99.8) | 99.8(20.3-99.8) | 99.8(96.7-99.8) |
| Calavi | 67567 | 99.9(43.0-100.0) | 100.0(96.2-100.0) | All | 99.3(16.3-99.9) | 100.0(52.4-100.0) | All |
| Glo Djibe | 18534 | 99.8(50.0-99.9) | 99.9(94.3-99.9) | All | 38.1(0.0-99.9) | 99.8(2.2-99.9) | 99.9(75.4-99.9) |
| Godomey | 100747 | 99.6(55.6-99.6) | All | All | 25.5(0.0-99.5) | 99.5(0.0-99.6) | 99.6(39.6-99.6) |
| Hèvié | 35721 | 99.6(40.3-99.9) | 99.9(88.2-99.9) | All | 0.0(0.0-93.7) | 52.9(0.0-99.9) | 99.9(0.0-99.9) |
| Kpanroun | 3754 | 97.9(40.4-99.7) | 99.7(98.0-99.7) | All | 18.8(0.0-97.9) | 99.5(0.0-99.7) | 99.7(62.8-99.7) |
| Ouèdo | 14992 | 100.0(52.0-100.0) | 100.0(91.5-100.0) | All | 0.0(0.0-100.0) | 100.0(0.0-100.0) | 100.0(1.5-100.0) |
| Togba | 34267 | 100.0(44.2-100.0) | 100.0(84.3-100.0) | All | 71.0(0.0-100.0) | 100.0(9.3-100.0) | 100.0(88.7-100.0) |
| Zinvie | 8773 | 98.6(77.4-99.6) | 99.6(98.5-99.6) | All | 98.5(38.3-99.6) | 99.6(78.3-99.6) | All |
| **ABOMEY-CALAVI** | **328331** | **99.7(46.5-99.8)** | **99.8(93.8-99.8)** | All | **51.3(4.8-99.1)** | **94.7(16.7-99.8)** | **99.8(62.5-99.8)** |
| 1er arr. | 17231 | 99.9(53.6-99.9) | All | All | 99.9(32.1-99.9) | 99.9(90.2-99.9) | All |
| 2eme arr. | 10016 | All | All | All | 99.9(96.5-99.9) | All | All |
| 3eme arr. | 11849 | 98.2(98.2-98.2) | 98.2(98.2-98.2) | 98.2(98.2-98.2) | 98.2(39.9-98.2) | 98.2(98.2-98.2) | 98.2(98.2-98.2) |
| 4eme arr. | 11309 | 98.5(91.2-98.5) | 98.5(98.5-98.5) | 98.5(98.5-98.5) | 98.5(80.2-98.5) | 98.5(98.5-98.5) | 98.5(98.5-98.5) |
| 5eme arr. | 11354 | All | All | All | All | All | All |
| 6eme arr. | 14074 | 99.9(98.0-99.9) | All | All | 99.9(67.5-99.9) | All | All |
| 7eme arr. | 9762 | All | All | All | 100.0(82.9-100.0) | All | All |
| 8eme arr. | 10669 | All | All | All | 100.0(97.2-100.0) | All | All |
| 9eme arr. | 15257 | 99.1(93.5-99.2) | All | All | 99.1(20.7-99.2) | 99.2(79.1-99.3) | All |
| 10eme arr. | 11404 | 100.0(100.0-100.0) | All | All | 100.0(65.0-100.0) | All | All |
| 11eme arr. | 9784 | All | All | All | 100.0(73.1-100.0) | All | All |
| 12eme arr. | 30348 | 99.8(87.4-99.8) | All | All | 92.0(38.9-99.8) | 99.8(60.0-99.8) | 99.8(98.8-99.8) |
| 13eme arr. | 23289 | 100.0(96.0-100.0) | All | All | All | 100.0(48.7-100.0) | All |
| **COTONOU** | **186346** | **99.7(91.8-99.7)** | All | All | **98.4(52.5-99.7)** | **99.7(84.2-99.7)** | All |
| 1 er arr. | 1392 | 99.8(11.6-100.0) | All | All | 94.0(0.0-100.0) | 100.0(0.0-100.0) | All |
| 2 eme arr. | 10028 | 100.0(93.3-100.0) | All | All | 99.8(32.4-100.0) | 100.0(96.1-100.0) | All |
| 3 eme arr. | 6772 | 100.0(67.1-100.0) | All | All | 99.4(0.0-100.0) | 100.0(86.9-100.0) | All |
| 4 eme arr. | 3091 | 99.7(68.6-99.7) | All | All | 99.3(0.0-99.7) | 99.7(29.0-99.7) | All |
| Avélékété | 2644 | 89.9(49.3-92.6) | 92.9(80.2-92.9) | 93.6(93.0-93.9) | 0.0(0.0-42.7) | 23.9(0.0-92.9) | 92.9(0.0-93.7) |
| Bossito | 3352 | 99.1(32.3-99.8) | 99.8(87.0-99.8) | All | 0.0(0.0-70.8) | 63.8(0.0-99.8) | 99.8(14.8-99.8) |
| Djegbadji | 2614 | 80.6(33.8-83.9) | 85.0(78.9-85.8) | 87.5(86.0-88.8) | 3.9(0.0-68.7) | 64.7(0.0-85.0) | 86.6(35.7-88.0) |
| Houakpe_ Daho | 989 | 62.2(40.5-80.3) | 82.6(54.6-84.9) | 88.2(78.2-89.6) | 0.0(0.0-63.4) | 62.1(0.0-83.3) | 86.4(51.7-89.0) |
| Pahou | 36918 | 99.3(42.3-99.5) | 99.5(96.4-99.5) | All | 0.4(0.0-65.4) | 50.9(0.0-99.5) | 99.5(1.5-99.5) |
| Savi | 9629 | 99.2(53.4-99.7) | 99.7(96.7-99.7) | All | 37.3(8.0-99.7) | 99.7(17.3-99.7) | 99.7(87.5-99.7) |
| **OUIDAH** | **77429** | **98.0(52.4-98.6)** | **98.7(95.3-98.8)** | **98.9(98.7-99.0)** | **32.2(5.2-78.7)** | **70.7(23.4-98.7)** | **98.8(41.6-99.0)** |
| 1e Arr. | 26461 | 99.9(71.3-99.9) | 99.9(99.3-99.9) | All | 99.6(11.6-99.9) | 99.9(69.6-99.9) | All |
| 2e Arr. | 6031 | 98.5(98.5-98.5) | 98.5(98.5-98.5) | 98.5(98.5-98.5) | 98.5(98.5-98.5) | 98.5(98.5-98.5) | 98.5(98.5-98.5) |
| 3e Arr. | 6595 | All | All | All | All | All | All |
| 4e Arr. | 22726 | 100.0(89.5-100.0) | All | All | 100.0(18.1-100.0) | 100.0(88.2-100.0) | All |
| 5e Arr. | 16114 | 100.0(92.9-100.0) | All | All | 100.0(45.4-100.0) | All | All |
| **PORTO-NOVO** | **77927** | **99.8(85.6-99.8)** | **99.8(99.6-99.8)** | All | **99.7(34.6-99.8)** | **99.8(86.1-99.8)** | All |
| Aglangandan | 22190 | 99.2(74.9-99.2) | All | All | 94.7(0.7-99.2) | 99.2(25.7-99.3) | 99.3(97.7-99.3) |
| Aholouyeme | 3355 | 99.2(39.3-99.3) | All | All | 12.4(0.0-98.7) | 99.0(0.0-99.5) | 99.5(73.5-99.7) |
| Djeregbe | 13805 | 99.8(64.8-99.8) | All | All | 98.4(2.8-99.8) | 99.8(45.1-99.8) | All |
| Ekpè | 28219 | 99.3(72.8-99.4) | All | All | 7.0(0.0-99.3) | 99.3(0.0-99.4) | 99.4(35.9-99.4) |
| SEmE - Kpodji | 12817 | 99.8(58.3-99.8) | All | All | 0.1(0.0-99.8) | 99.8(0.0-99.8) | 99.8(22.1-99.8) |
| Tohoue | 19055 | 99.6(54.0-99.7) | All | All | 1.8(0.0-91.1) | 77.1(0.0-99.7) | 99.7(27.5-99.7) |
| **SEME PODJI** | **99441** | **99.5(65.6-99.5)** | **99.5(98.3-99.5)** | All | **37.6(0.6-97.8)** | **95.2(12.0-99.5)** | **99.5(56.4-99.6)** |

4 Monitoring emergency obstetric care a handbook.

5 Transforming sanitation in Benin: A tale of resilience and renewal. https://blogs.worldbank.org/en/water/transforming-sanitation-benin-tale-resilience-and-renewal (accessed 15 July 2024)

6 Geofabrik GmbH. Download OpenStreetMap data for this region: Benin. 2024. https://download.geofabrik.de/africa/benin.html (accessed 20 September 2024)

7 Impact Observatory Inc. Methodology & Accuracy Summary: 10m Global Land Use Land Cover Maps. Published Online First: July 2022.

8 Karra K, Kontgis C, Statman-Weil Z, *et al.* Global land use/land cover with Sentinel-2 and deep learning. *IGARSS 2021-2021 IEEE International Geoscience and Remote Sensing Symposium*. Brussels: IEEE 2021.

9 Esri. ArcGIS Living Atlas of the World. Published Online First: 2024.

10 Van Zyl JJ. The shuttle radar topography mission (SRTM): A breakthrough in remote sensing of topography. *Acta Astronautica*. 2001.

11 Tobler W. Three presentations on geographical analysis and modeling: Non-isotropic geographic modeling speculations on the geometry of geography global spatial analysis. *Technical report (National center for geographic information and analysis)*. 1993.

12 Chi G, Fang H, Chatterjee S, *et al.* Microestimates of wealth for all low- and middle-income countries. *Proc Natl Acad Sci U S A*. 2022;119. doi: 10.1073/pnas.2113658119

13 HDX. Relative Wealth Index. 2021. https://data.humdata.org/dataset/relative-wealth-index? (accessed 5 July 2024)

14 Dijkstra L, Florczyk AJ, Freire S, *et al.* Applying the Degree of Urbanisation to the globe: A new harmonised definition reveals a different picture of global urbanisation. *J Urban Econ*. 2021;125. doi: 10.1016/j.jue.2020.103312

15 Schiavina Marcello, Melchiorri Michele, Pesaresi Martino. GHS-SMOD R2023A - GHS settlement layers, application of the Degree of Urbanisation methodology (stage I) to GHS-POP R2023A and GHS-BUILT-S R2023A, multitemporal (1975-2030). 2023.

16 Tatem AJ. WorldPop, open data for spatial demography. Sci Data. 2017;4.

17 Pezzulo C, Hornby GM, Sorichetta A, *et al.* Sub-national mapping of population pyramids and dependency ratios in Africa and Asia. *Sci Data*. 2017;4. doi: 10.1038/sdata.2017.89

18 U.S. Census Bureau. Benin Annual Five-Year Age Group Population Estimates by Sex for 2015 to 2030: National, and First- and Second-Order Administrative Divisions . 2020. https://www2.census.gov/programs-surveys/international-programs/tables/time-series/pepfar/benin.xlsx (accessed 3 October 2024)

19 Mathias Grovogui F, Kpogomou P, Mina Millimouno T, *et al.* Geographical accessibility and inequalities in access to childbirth care in the Grand Conakry metropolitan area, Guinea: a spatial modelling study. *Research Square (preprint)*. Published Online First: 16 September 2024. doi: 10.21203/rs.3.rs-4926298/v1

20 Macharia PM, Banke-Thomas A, Beňová L. Advancing the frontiers of geographic accessibility to healthcare services. *Communications Medicine*. 2023;3:158. doi: 10.1038/s43856-023-00391-w

21 Kobo. The KoboToolbox software. 2024. https://www.kobotoolbox.org/about-us/software/ (accessed 12 August 2024)

22 Ray N, Ebener S. AccessMod 3.0: Computing geographic coverage and accessibility to health care services using anisotropic movement of patients. *Int J Health Geogr*. 2008;7. doi: 10.1186/1476-072X-7-63

23 University of Geneva/GeoHealth group, World Health Organization, MORU/Health GeoLab Group. AccessMod 5. 2023. https://www.accessmod.org/ (accessed 25 May 2023)
